# Short-Term Mortality After Opioid Initiation Among Opioid-Naïve and Non-Naïve Patients with Dementia: A Retrospective Cohort Study

**DOI:** 10.1101/2024.11.25.24317747

**Authors:** Yeon-Mi Hwang, Jennifer M. Hah, Jennifer E. Bramen, Jennifer J. Hadlock, Tina Hernandez-Boussard

## Abstract

**Background:** Despite the ongoing opioid epidemic, the mortality risk of opioid initiation in patients with dementia or mild cognitive impairment (MCI) remains understudied despite their vulnerability. This study evaluates mortality risks associated with opioid use in patients diagnosed with dementia or MCI by comparing outcomes between new and consistent users.

**Methods:** We conducted a retrospective cohort study using data from Northern California academic healthcare system (2015/01/01-2024/07/31), including 27,757 patients aged 50–100 with dementia or MCI. Of these, 14,105 received opioids after diagnosis and were classified as new (opioid-naïve; n=9444) or consistent (non-naïve; n=4663) users. Cox regression assessed 14-day mortality risk. Aalen’s additive model examined time-varying impact. Causes of death were extracted from clinical notes using GPT-3·5-Turbo. Findings were validated in community healthcare system cohort (n=207,873) across western U.S. states (2015/01/01-2023/05/31).

**Findings:** In the primary cohort, 4.1% (572/14105) of patients died within 14 days of opioid exposure. New users had a significantly higher 14-day mortality risk than consistent users (adjusted hazard ratio [aHR], 2·00 [1·59–2·52]; P<0·0001). The validation cohort had a 14-day mortality rate of 6·2% (7022/113343) with a smaller difference between new (n=77,204) and consistent (n=36,194) users (aHR 1·20 [1·13–1·27]; P<0·0001). In both cohorts, elevated risk stabilized after day 30. In the primary cohort, respiratory conditions, particularly pneumonia, were more prevalent among new users who died early.

**Interpretation:** Opioid initiation in these patients is associated with increased short-term mortality in new users, underscoring the need for cautious initiation and close monitoring during the first month.

**Funding:** NIH

## 1. Introduction

Pain is a common experience among patients with dementia,^1–3^ frequently arising from age-related comorbidities including arthritis, fractures, and other chronic conditions.^4–6^ Managing pain in this population is particularly challenging as cognitive impairment interferes with accurate pain assessment.^2^ While opioids are commonly prescribed for moderate to severe pain, they carry substantial risks in the elderly population, including respiratory depression, falls, sedation, and confusion.^7^ These risks are particularly concerning at the start of therapy, due to lower tolerance and increased overdose risk.^8–10^ For dementia patients, who often struggle to communicate pain symptoms, the potential for adverse outcomes may be even greater.^2^

Given their vulnerability, several studies have examined the incident opioid use in this group. Taipale et al.^11^ reported an elevated risk of hip fracture among new opioid users, with an adjusted hazard ratio (aHR) of 1·96 [1·27-3·02] in the first two months. Hamina et al.^12^ observed a higher risk of hospital-treated pneumonia in dementia patients exposed to opioids. More recently, Jensen-Dahm et al.^13^ found a significantly higher mortality risk among opioid-exposed dementia patients within 180 days (aHR, 4·13 [3·98–4·30]), with a peak risk in the first 14 days (aHR 10·95 [9·87–12·15]).

Previous studies primarily compared opioid-exposed and unexposed patients, potentially introducing confounding by indication. Instead, our study evaluates short-term mortality by comparing patients who initiated opioids after dementia onset with those who had ongoing opioid exposure from the year preceding diagnosis, aiming to isolate the impact of opioid initiation. We hypothesized that opioid-naïve patients, those newly exposed post-diagnosis, would experience higher mortality rates due to reduced tolerance to adverse effects. We validated our findings from a primary cohort of academic hospitals in a larger community healthcare system spanning western U.S states. Additionally, we employed a large language model (LLM) in the primary cohort to analyze unstructured clinical notes, enabling detailed characterization of conditions present at death.

## 2. Methods

### 2.1 Study design and participants

This retrospective cohort study was primarily conducted at Stanford Health Care Alliance (SHCA), a large academic healthcare system. The study was approved by the Stanford University Institutional Review Board (Protocol 47644). We validated our findings using data from Providence St. Joseph Health (PSJH), a large, not-for-profit integrated U.S. community healthcare system serving both urban and rural populations across seven states (Alaska, California, Montana, Oregon, New Mexico, Texas, and Washington), with most care sites concentrated in the western U.S. This validation study was approved by the PSJH Institutional Review Board (Protocol STUDY2024000402).

Cohort selection of the primary cohort is described in Figure 1. From individuals with encounters at SHCA between 2015/01/01 and 2024/07/31 (n = 3,247,002), we identified those with at least one diagnosis of dementia or mild cognitive impairment (MCI) (n = 32,460; Table S1). We included MCI due to potential overlap with early-stage dementia. The cohort was limited to individuals aged 50 to 100 at first diagnosis (n = 31,252) and excluded those who did not survive beyond 14 days post-surgery (n = 31,187), as opioids are routinely administered perioperatively, making it difficult to distinguish whether early deaths were due to surgical complications or opioid. To ensure continuity of care, we included only individuals with at least three encounters both before and after their first diagnosis and those who survived more than 7 days after their initial dementia/MCI diagnosis (n = 27,757).

**Figure 1.**
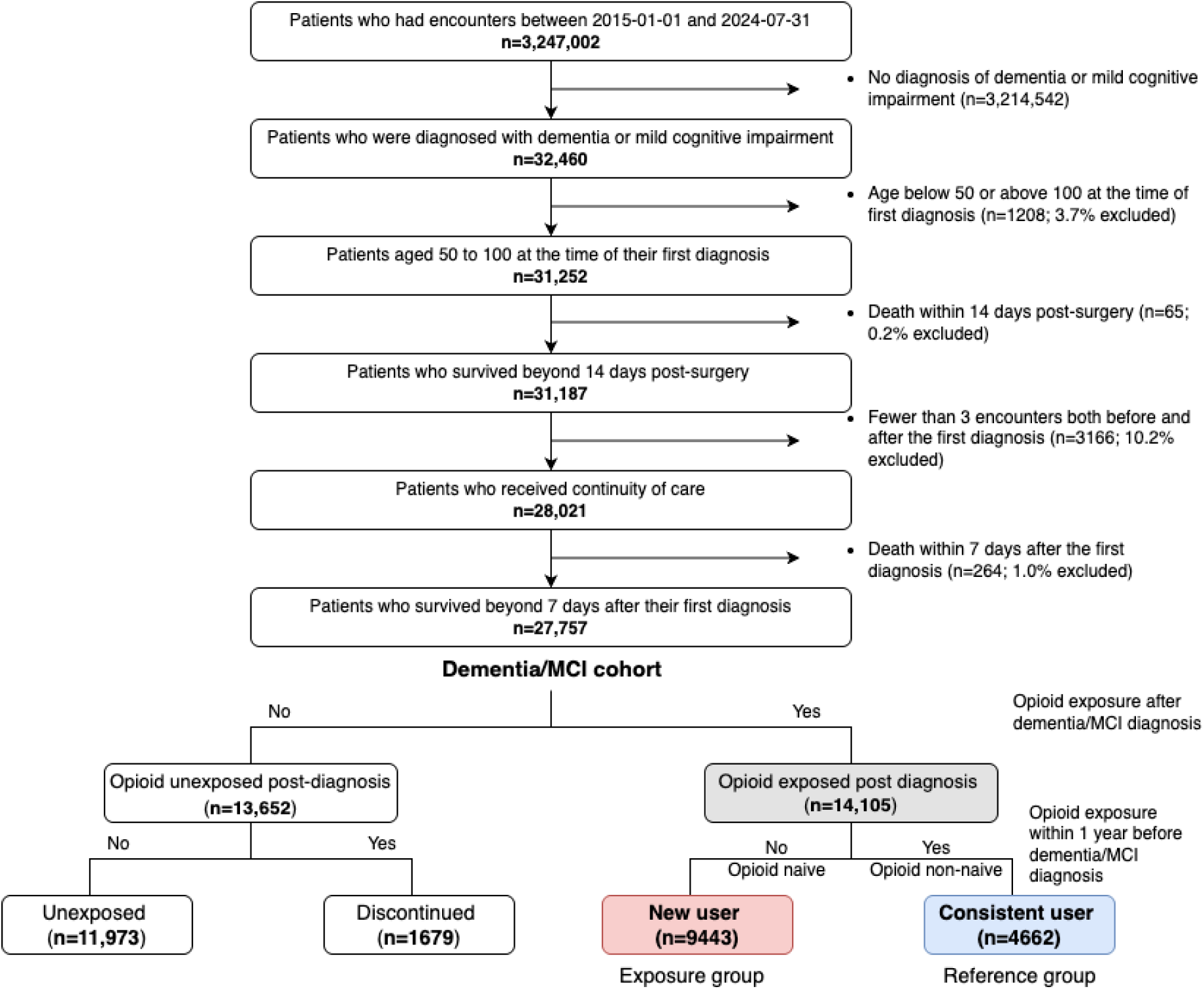
Flowchart of primary cohort and exposure group selection. Descriptive statistics of the dementia/MCI cohort comparing opioid-exposed and unexposed groups are presented in Table S8. Descriptive statistics of the opioid-exposed cohort comparing new users and consistent users are presented in Table 1 and Table S7.

**Table 1.** Descriptive statistics of the analytic cohort. Categorical variables were compared using the chi-squared test. Continuous variables were evaluated using Welch’s t-test. The Benjamini-Hochberg method was used to correct for multiple testing errors. All analyses were conducted using the PyPI package tableone (v·0·9·1)^26^. Variables that contained cells with fewer than 20 cases were dropped to protect patient privacy. Variables are defined in Table S3.

|  | Missing | Overall<br>(n=14,107) | new user (n=9444) | consistent user<br>(n=4663) | P |
| --- | --- | --- | --- | --- | --- |
| **Dementia category** |  |  |  |  |  |
| Mild cognitive impairment, n (%) |  | 5213 (37.0) | 3182 (33.7) | 2031 (43.6) | <0.001 |
| Alzheimer's disease, n (%) |  | 3160 (22.4) | 2411 (25.5) | 749 (16.1) | <0.001 |
| Vascular dementia, n (%) |  | 1319 (9.3) | 881 (9.3) | 438 (9.4) | 0.93 |
| Other/Unspecified dementia, n (%) |  | 10637 (75.4) | 7390 (78.3) | 3247 (69.6) | <0.001 |
| **Patient characteristics** |  |  |  |  |  |
| Sex | 1 |  |  |  | 1.000 |
| Female |  | 7904 (56.0) | 5292 (56.0) | 2612 (56.0) |  |
| Male |  | 6202 (44.0) | 4152 (44.0) | 2050 (44.0) |  |
| Age at diagnosis, median [Q1,Q3] | 0 | 81.0 [73.0,87.0] | 81.0 [74.0,87.0] | 79.0 [71.0,86.0] | <0.001 |
| Age group, n (%) | 0 |  |  |  | <0.001 |
| 50-59 |  | 695 (4.9) | 398 (4.2) | 297 (6.4) |  |
| 60-69 |  | 1720 (12.2) | 986 (10.4) | 734 (15.7) |  |
| 70-79 |  | 4080 (28.9) | 2643 (28.0) | 1437 (30.8) |  |
| 80-89 |  | 5499 (39.0) | 3856 (40.8) | 1643 (35.2) |  |
| 90-100 |  | 2113 (15.0) | 1561 (16.5) | 552 (11.8) |  |
| Race, n (%) | 57 |  |  |  | <0.001 |
| White |  | 9021 (64.2) | 6007 (63.8) | 3014 (65.0) |  |
| Black |  | 2019 (14.4) | 1433 (15.2) | 586 (12.6) |  |
| Asian |  | 725 (5.2) | 422 (4.5) | 303 (6.5) |  |
| Native American/Hawaiian/Pacific Islander |  | 117 (0.8) | 73 (0.8) | 44 (0.9) |  |
| Unknown/Refused |  | 265 (1.9) | 209 (2.2) | 56 (1.2) |  |
| Other |  | 1903 (13.5) | 1266 (13.5) | 637 (13.7) |  |
| Ethnicity, n (%) | 13 |  |  |  | <0.001 |
| Hispanic/Latino |  | 1326 (9.4) | 835 (8.9) | 491 (10.5) |  |
| Non-Hispanic/Latino |  | 12402 (88.0) | 8326 (88.2) | 4076 (87.5) |  |
| Unknown/Refused |  | 366 (2.6) | 274 (2.9) | 92 (2.0) |  |
| Insurance, n (%) | 226 |  |  |  | 0.78 |
| Medicare/Medicaid |  | 7402 (53.3) | 4932 (53.2) | 2470 (53.6) |  |
| Private |  | 4481 (32.3) | 3013 (32.5) | 1468 (31.9) |  |
| Other |  | 1998 (14.4) | 1332 (14.4) | 666 (14.5) |  |
| **BMI, median [Q1,Q3]** | 914 | 25.4 [22.3,28.9] | 25.1 [22.1,28.6] | 25.8 [22.6,29.6] | <0.001 |
| **BMI category, n (%)** | 1158 |  |  |  | <0.001 |
| Under weight |  | 624 (4.8) | 426 (5.0) | 198 (4.5) |  |
| Normal weight |  | 5436 (42.0) | 3717 (43.5) | 1719 (39.1) |  |
| Over weight |  | 4451 (34.4) | 2913 (34.1) | 1538 (34.9) |  |
| Obese |  | 1812 (14.0) | 1127 (13.2) | 685 (15.6) |  |
| Severely obese |  | 626 (4.8) | 365 (4.3) | 261 (5.9) |  |
| **Death outcome** |  |  |  |  |  |
| **Death, n (%)** | 0 | 6104 (43.3) | 4140 (43.8) | 1964 (42.1) | 0.06 |
| **Days to death from first opioid exposure, mean (SD)** | 8004 | 837.4 (910.6) | 837.7 (913.9) | 836.8 (904.0) | 0.97 |
| **Death within 14 days after first opioid exposure, n (%)** | 0 | 572 (4.1) | 451 (4.8) | 121 (2.6) | <0.001 |
| **Death within 60 days after first opioid exposure, n (%)** | 0 | 1261 (8.9) | 882 (9.3) | 379 (8.1) | 0.02 |
| **Death within 180 days after first opioid exposure, n (%)** | 0 | 1997 (14.2) | 1379 (14.6) | 618 (13.3) | 0.03 |

The same inclusion criteria was applied to the PSJH validation cohort during an observation period 2015/01/01-2023/05/31, yielding 207,873 patients (detailed cohort selection is in Figure S1)

### 2.2 Measures

#### 2.2.1 Outcome

Our primary outcome was short-term mortality, defined as death within 14 days after the first opioid exposure; with additional follow-up to 180 days. Death dates were obtained from the EHR and state’s public records. While the EHR provides comprehensive longitudinal health profiles from both structured fields and clinical notes, deaths occurring outside the healthcare system may be missed. Therefore, we incorporated statewide death records to capture out-of-hospital deaths. Although deaths among patients residing outside their state may not be captured, we anticipate minimal impact on our findings given our cohort was restricted to elders receiving continuous care within the system.

#### 2.2.2 Exposure

Our exposure of interest was opioid initiation following dementia/MCI onset (Figure 1 and S1). To isolate the impact of opioid initiation and minimize the confounding by indication, we classified post-diagnosis opioid-exposed patients into two groups based on their opioid use in the year preceding dementia/MCI onset: opioid-naïve (new users) and non-naïve (consistent users). The exposure group (new users) comprised individuals who received their first opioid prescription after the onset of dementia/MCI, with no opioid use in the preceding year. The reference group (consistent users) included individuals who used opioids within one year before their initial dementia or MCI diagnosis and continued use afterward. Opioid medications included were buprenorphine, codeine, fentanyl, hydrocodone, hydromorphone, meperidine, methadone, morphine, oxycodone, and tramadol (Table S2). The observation period began at the start date of the first opioid use after dementia/MCI onset.

#### 2.2.3 Covariates

Patient characteristics and health conditions were collected from EHR prior to the first opioid exposure including age, race, ethnicity, body mass index (BMI), insurance status, comorbidities, and medication (variable definitions in Table S3).

We included comorbidities associated with dementia and mortality, from the two years before first opioid exposure, spanning cardiovascular, respiratory, metabolic, liver, renal, cancer, neurological, mental health and other conditions (complete list with ICD codes in Table S4).

We examined medication use within one year before first opioid exposure, focusing on anti-dementia drugs,^14^ medications in American Geriatrics Society’s (AGS) Beers Criteria.^15^ and clinician-recommended (JMH) dementia-relevant medications. Medications were grouped medications into therapeutic/chemical subcategories (complete list with RxNorm codes in Table S5).

### 2.3 Analyses

#### 2.3.1 Descriptive analysis

We compared covariates between new and consistent users using chi-squared tests for categorical variables and Welch’s t-test for continuous variables, with Benjamini-Hochberg correction. We also performed descriptive analysis of opioid-exposed versus unexposed patients.

#### 2.3.2 Statistical analysis

##### 2.3.2.1 Main analysis

We employed Cox proportional hazards model to compute 14-day mortality hazard ratios (HR) using lifelines PyPI (v 0·29·0),^16^ with follow-up to 180 days. Model performance was assessed with Concordance Index (C-Index). C-index ranges from 0·5 to 1, where 1 indicates perfect prediction. Time-varying exposure effects were analyzed using Aalen’s additive hazards model, evaluated with Integrated Brier Score (IBS). IBS ranges from 0 to 1. IBS of 0 indicates perfect accuracy. We identified stabilization points where the smoothed first derivative (10-day rolling mean) of cumulative hazard remained below 5% of its maximum for 10 consecutive days. Variables with <5% cases were excluded, and missing values were imputed using medians. Analysis was limited to 180 days as we assumed the impact of opioid initiation diminishes over time and increasing confounding from other factors.

##### 2.3.2.2 Subgroup, sensitivity, post-hoc supplementary analysis

For sensitivity analyses, we compared first-time users with long-term consistent users (≥90 days pre-diagnosis opioid use) and tested the proportional hazard assumption using Schoenfeld residuals and the Kolmogorov-type supremum test. We conducted subgroup analyses by opioid strength (weak and strong)^17^, dementia/MCI status, which are not mutually exclusive, and medication order mode (inpatient and outpatient). Based on analysis of unstructured notes, we examined pneumonia’s role by analyzing patients who had a pneumonia diagnoses within 7 days prior to opioid initiation, and by comparing pneumonia risk between new and consistent users among those without prior pneumonia.

#### 2.3.3 Identification of potential reasons for short-term mortality

To examine causes of death within 14 days of first opioid exposure, we securely processed clinical notes from the three days preceding death using GPT-3·5 Turbo to identify top three contributing conditions in the primary cohort (Table S6).^18^ This was done to uncover clinical correlations not readily apparent in structured data alone. We validated GPT outputs by examining 50 random notes for hallucinations errors and comparing another 50 notes with explicit death diagnoses. Health conditions were categorized into predefined categories using GPT-4o with clinician (JMH) review of mappings. Multiple assignment was allowed.

### 2.4 Validation

Variables were collected using similar methods as the primary cohort, adapted for different data models. The same analyses were performed, except for GPT analysis.

## 3. Results

Among 27,757 individuals with dementia/MCI, 14,105 (50·8%) received opioids post-diagnosis: 9443 new users and 4662 consistent users (Figure 1). The opioid-exposed cohort was predominantly female (56·0%), White (64·2%), non-Hispanic (88·0%), Medicaid/Medicare insured (53·3%), with normal BMI (42·0%), and median age of 81 years (IQR: 73–87). Mortality rates were 4·1% at 14 days, 8·9% at 60 days, and 14·2% at 180 days (Figure 1 and Table 1, S7, S8).

Compared to consistent users, new users were older, had more Asian and fewer Black/Hispanic patients, less obesity, higher dementia and lower MCI prevalence (P<0·001, Table 1). They had fewer comorbidities (33/34 conditions) and medications (43/45; P<0·05). Mortality was higher in new users at 14 days (4·8% vs. 2·6%, P<0·001), 60 days (9·3% vs. 8·1%, P<0·05), and 180 days (14·6% vs. 13·3%, P<0·05; Table S1, S7).

The validation cohort is described in Table S9. Of 207,873 individuals with dementia or MCI, 112,876 received opioids post-diagnosis (76,770 new users, 36,106 consistent users). Compared to the primary cohort, the validation cohort had more dementia (85·8% vs. 78·4%; P<0·0001) and less MCI diagnoses (26·9% vs. 36·9%; P<0·0001), higher proportions of female (60·2% vs. 56·0%; P<0·0001), White (82·2% vs. 64·2%; P<0·0001), non-Hispanic (91.2% vs. 88%; P<0·0001), and Medicare/Medicaid (92·7% vs. 53·3%; P<0·0001) patients. Overall mortality rates were higher in the validation cohort compared to the primary cohort (14-day: 5·6% vs. 4·0%, 60-day: 10·3% vs. 9·1%, 180-day:14·9%; P<0·0001). The patterns observed in the primary cohort, differences between new and consistent users in patient characteristics, comorbidities, medication exposures, and mortality, were similarly observed in the validation cohort.

In the primary cohort, new users showed significantly higher 14-day mortality risk, with an unadjusted HR of 1·86 [1·53, 2·28] increasing to 2·00 [1·59, 2·52] after adjustment (P<0·0001; C-Index: 0·78; Figure 2). In the validation cohort, the adjusted HR was 1·23 [1·16–1·30] (P < 0·0001), compared to an unadjusted HR of 1·59 [1·50–1·68], with a C-Index of 0·85 (Figure 3). Over 180 days, both cohorts showed consistently higher hazards in new users (Figure S2), with time-varying cumulative hazard stabilizing around day 30 (IBS; primary: 0·08, validation: 0·09). The cumulative hazard continued to rise in the primary cohort but plateaued in the validation cohort.

**Figure 2.**
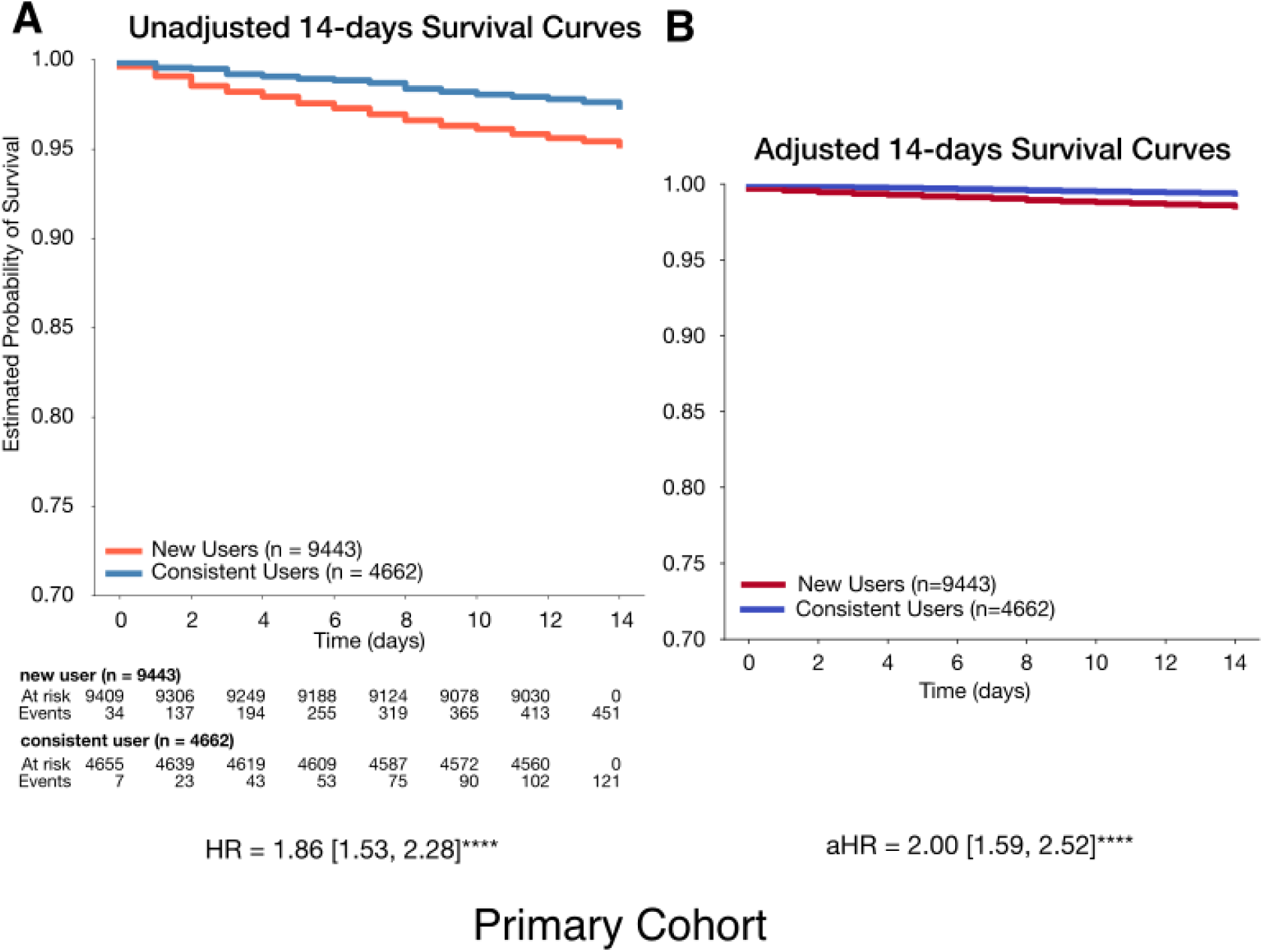
14-day survival probability curves comparing new users and consistent users of the primary cohort. A. Unadjusted 14-day survival probability curves comparing new users and consistent users B. Adjusted 14-day survival probability curves comparing new users and consistent users. New users (n=9443) had a lower likelihood of survival during the 14-day follow-up period, with an adjusted hazard ratio (aHR) of 2·00 [1·55, 2·47] (P<0·0001). The model achieved a concordance index of 0·78, indicating good predictive performance. Hazard ratios were computed using a Cox proportional hazards model, adjusted for age at diagnosis, race, ethnicity, body mass index (BMI), insurance status, comorbidities, and medication exposures. Complete lists of comorbidities and medications can be found in Tables S4 and S5. Variables with cases fewer than <5% of analytic cohort were excluded from the analysis. Significance levels: +: P < 0·1, *: P < 0·05, **: P < 0·01, ***: P < 0·001, ****: P < 0·0001.

**Figure 3.**
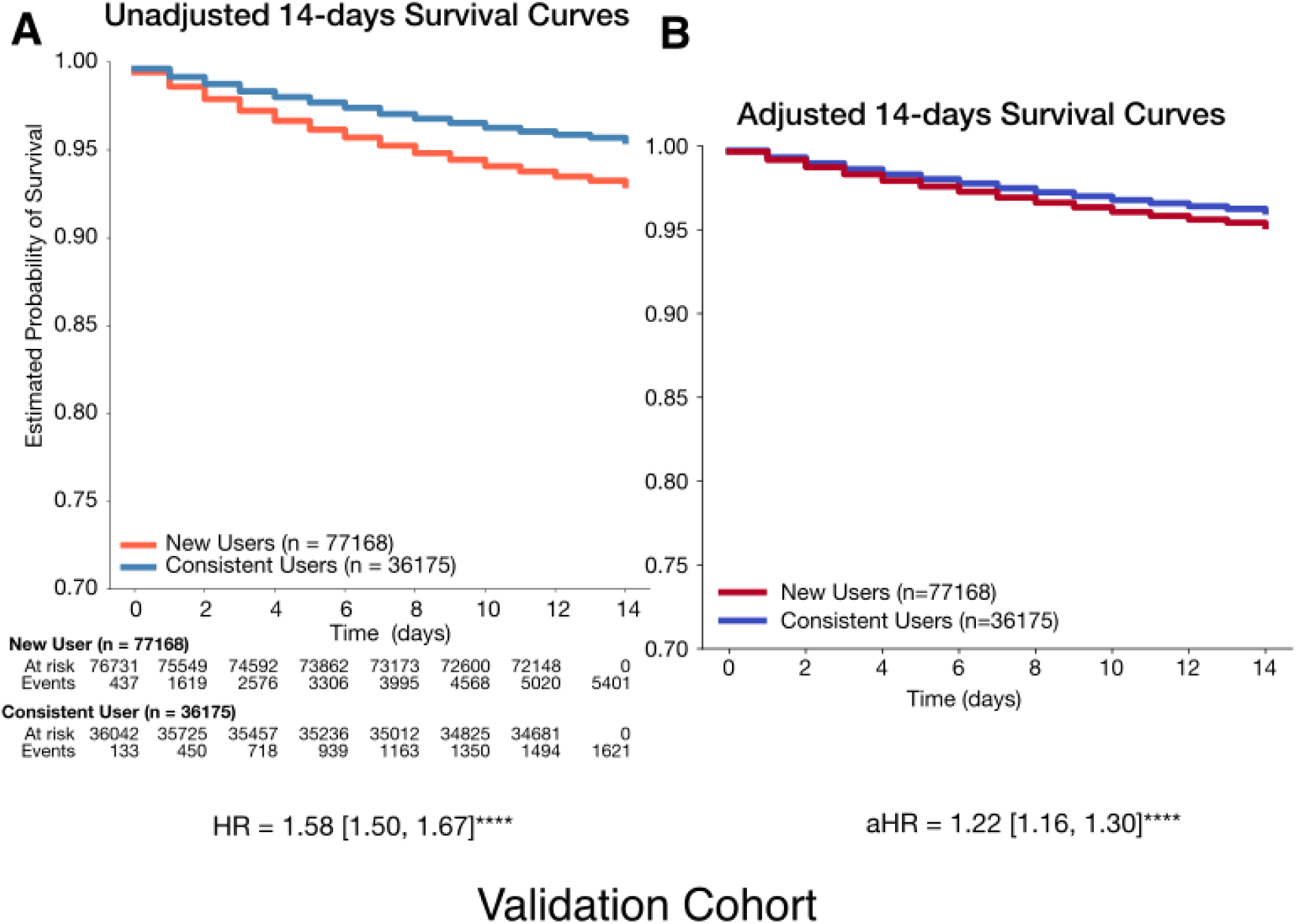
14-day survival probability curve comparing new users and consistent users of the valid. A. Unadjusted 14-day survival probability curves comparing new users and consistent users B. Adjusted 14-day survival probability curves comparing new users and consistent users. New users (n=77,168) had a lower likelihood of survival during the 14-day follow-up period, with an adjusted hazard ratio (aHR) of 1.22 [1.16, 1.30] (P<0·0001). The model achieved a concordance index of 0.85, indicating good predictive performance. Hazard ratios were computed using a Cox proportional hazards model, adjusted for age at diagnosis, race, ethnicity, body mass index (BMI), insurance status, comorbidities, and medication exposures. Complete lists of comorbidities and medications can be found in Tables S4 and S5. Variables with cases fewer than <5% of analytic cohort were excluded from the analysis. Significance levels: +: P < 0·1, *: P < 0·05, **: P < 0·01, ***: P < 0·001, ****: P < 0·0001.

Sensitivity analyses comparing new users to long-term consistent users showed similar survival trends and hazard ratio consistent with the main analysis in both cohorts (Figure S3). Accounting for variables that violated the proportional hazards assumption had minimal impact on the results.

Subgroup analyses consistently showed higher mortality risk for new users, with some differences between cohorts. In the primary cohort, mortality risk was higher with stronger opioids, in patients with dementia, and among inpatients. The validation cohort showed smaller hazard ratios overall, with larger hazard ratios in the MCI and inpatient groups, though dementia and outpatient groups had higher absolute mortality (Figure S4, S5, S6).

Within 14 days of their first opioid exposure post diagnosis, 450 new users and 121 consistent users died (Table 1). We processed 438 notes for 179 new users and 138 notes for 62 consistent users. No hallucinations were detected in a random 50 notes. In another random 50 notes with a specified principal diagnosis at death, 49 matched the specified principal diagnosis category (46 exact, 3 category match), and one partially matched (Table S10). Figure 5 shows that respiratory-related conditions exhibited the largest difference in prevalence between the groups, occurring in 62% of new users compared to 48% of consistent users (P=0.09). A more detailed breakdown of respiratory issues revealed that respiratory failure was the most common condition in both groups. However, pneumonia-related conditions were particularly high among new users, with a 19% higher prevalence (P=0·04).

**Figure 4.**
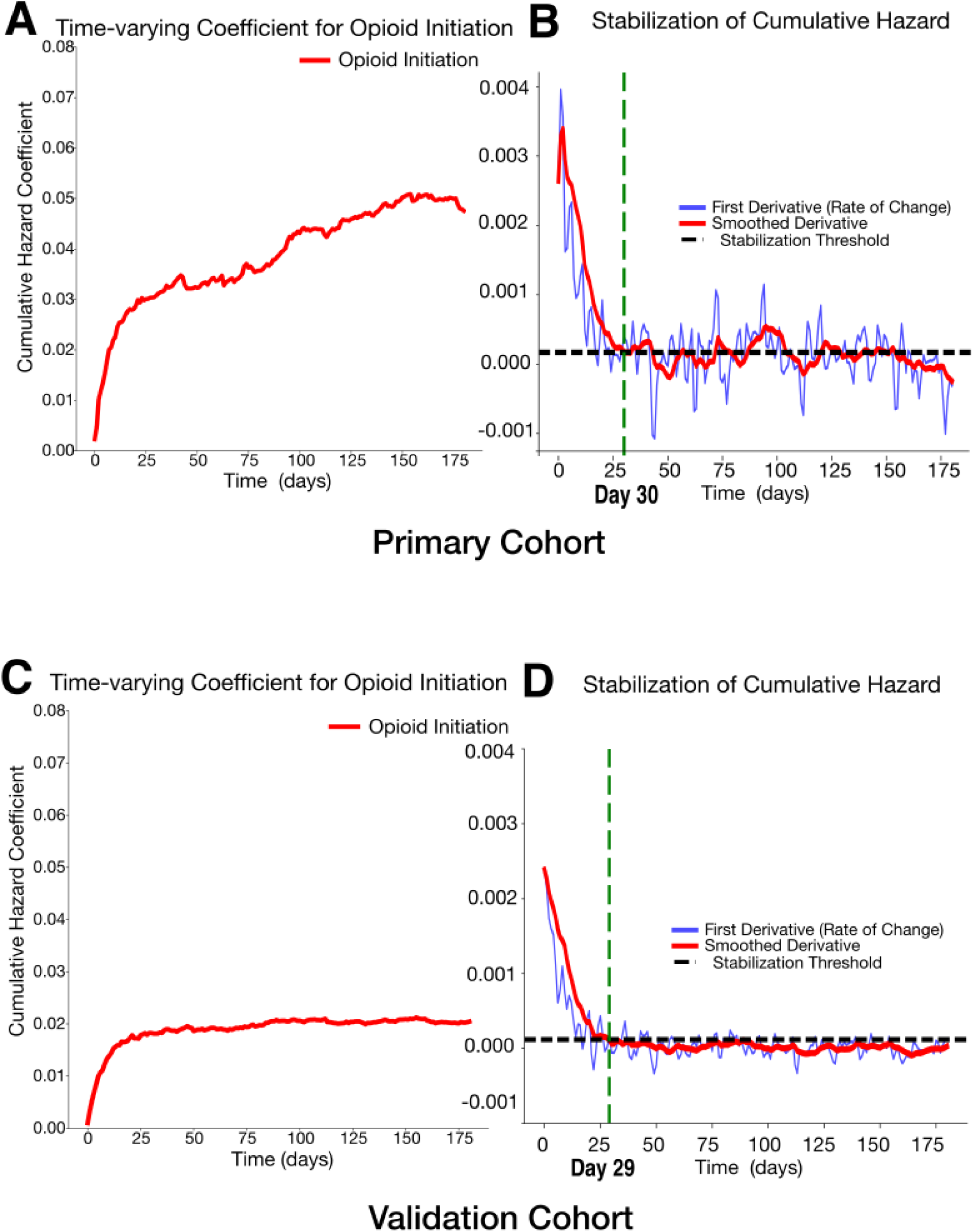
Time-varying effects of opioid initiation on cumulative hazard and stabilization. A. The cumulative hazard coefficient for opioid initiation over time in primary cohort (Integrated Brier Score [IBS]: 0.08, IBS of 0 means perfect accuracy, and IBS of 1 means perfect inaccuracy) B. The first derivative (rate of change) of the cumulative hazard (blue) and its smoothed version (red), with the stabilization threshold (black dashed line) in primary cohort C. The cumulative hazard coefficient for opioid initiation over time in validation cohort (IBS: 0.09) D. The first derivative (rate of change) of the cumulative hazard (blue) and its smoothed version (red), with the stabilization threshold (black dashed line) in valiation cohort. The green dashed line marks the estimated stabilization point (Day 30 for the primary cohort and Day 29 for the validation cohort), indicating when the cumulative hazard change stabilizes.

**Figure 5:**
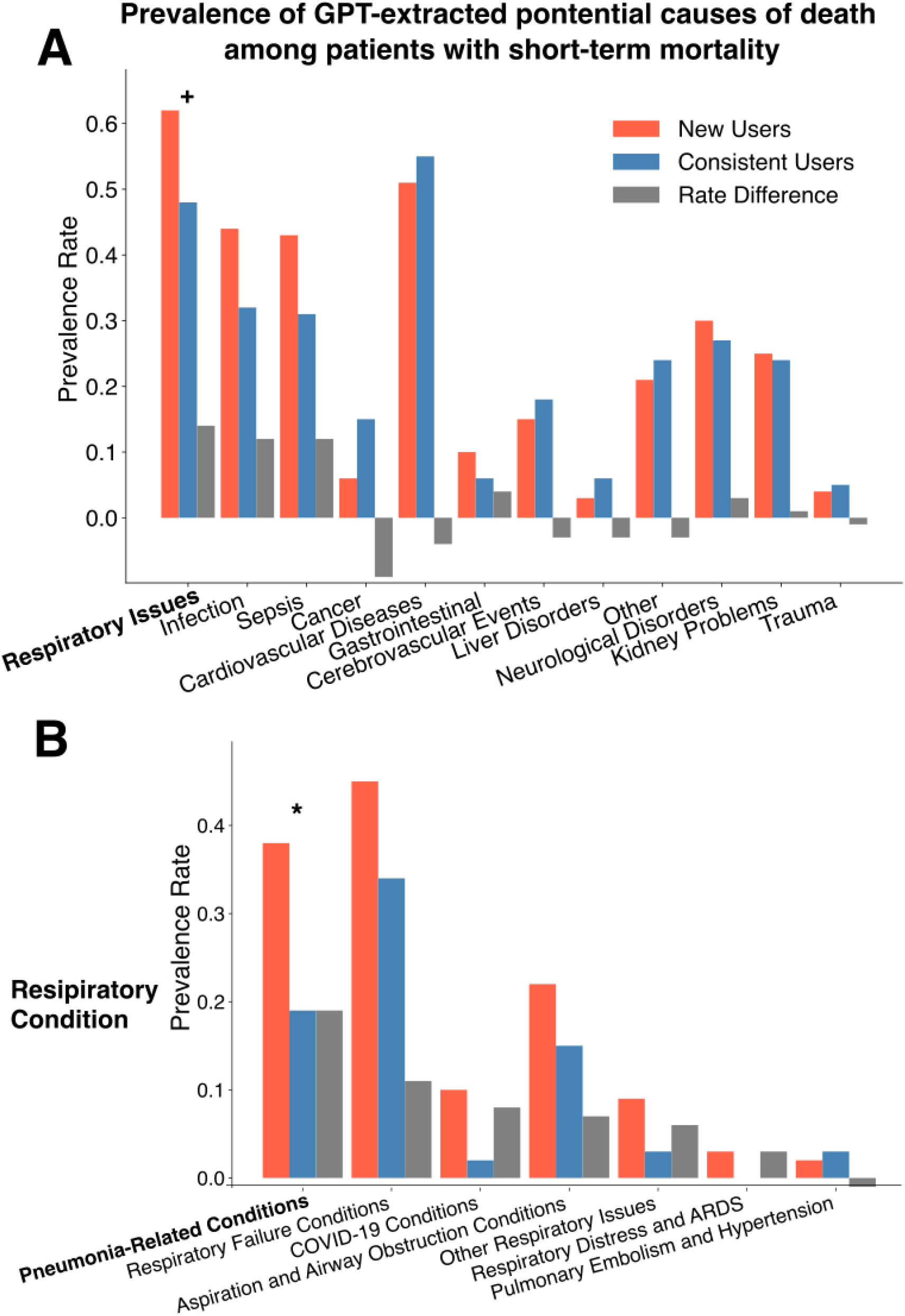
Bar plot of prevalence rate difference in GPT-extracted potential causes of death between new and consistent users in primary cohort. Plot A shows the difference in prevalence rates of GPT-extracted potential causes of death between new and consistent users, shown as a bar plot. Plot B shows the difference in prevalence rate focusing on specific respiratory-related conditions. We collected clinical notes mentioning death among individuals who passed away within 14 days after opioid initiation (438 notes for 179 patients in new users and 138 notes for 62 patients in consistent users). These notes were processed through secure GPT-3·5 Turbo to identify potential causes of death. The prompt used for this analysis is provided in Table S6. Among 50 randomly selected notes, no instances of hallucination were detected. If a note did not contain information about the health condition at the time of death, GPT-3·5 Turbo outputted ‘NA’. When a primary cause of death was specified, it was listed as the first cause; if unspecified, GPT-3·5 Turbo identified the primary health concern mentioned in the note at the time of death. We utilized GPT-4o to categorize these potential causes into predefined health condition categories. If a cause fit into multiple categories, it was assigned to all relevant categories. A clinician (JMH) reviewed the cause-category mappings to ensure accuracy, and adjustments were made based on their feedback. Significance levels: +: P < 0·1, *: P < 0·05, **: P < 0·01, ***: P < 0·001, ****: P < 0·0001.

Opioid initiation was associated with a higher mortality risk in patients with pre-existing pneumonia in the primary cohort (aHR = 2·45 [1·51, 3·96], P<0.001) but not in the validation cohort. For patients without pre-existing pneumonia, the validation cohort showed a slight but significant increase in pneumonia risk (aHR = 1·22 [1·08, 1·38], P<0·01), while the primary cohort did not (Figure S7).

## 4. Discussion

Initiating opioids in opioid-naïve patients with dementia or MCI was associated with increased 14-day mortality compared to those with prior opioid exposure. This association was consistent across both an academic medical center and a larger community healthcare system persisting through sensitivity and subgroup analyses. The heightened risk associated with opioid initiation stabilized by day 30 in both cohorts.

Our study is the first to compare short-term mortality between patients who newly initiated opioids after dementia diagnosis and those with prior and ongoing opioid exposure, reducing confounding by indication. A recent study from Denmark reported an 11-fold increase in 14-day mortality risk associated with opioid exposure among patients with dementia. Their analysis compared opioid-exposed to unexposed patients using a one-year washout period before dementia diagnosis.^13^ Instead, we focused within the opioid-exposed group, assuming underlying health conditions would be similar between the exposure group (new users) and reference group (consistent users). In our study, consistent users even had higher rates of comorbidities and medication use and new users still showed elevated mortality risk.

We found elevated 14-day mortality risk among new opioid users, with this association persisting across distinctly different healthcare settings. Both the Northern California academic medical center and the Western U.S. community hospital system showed increased mortality risk, as confirmed through comprehensive sensitivity and subgroup analyses. The consistency of this finding is particularly noteworthy given the distinct differences in healthcare delivery models and patient populations between these institutions. The validation cohort, serving both rural and urban communities through a not-for-profit system, had a predominantly government-insured population (93%) compared to the academic center’s more diverse payer mix (52%), reflecting substantial variations in patient characteristics and socioeconomic factors. Previous research has established that such institutional and demographic differences can significantly influence clinical outcomes.^19–21^ Interestingly, the same covariate adjustments increased the hazard ratio in the primary cohort but reduced it in the validation cohort. These findings strengthen the generalizability of our results while highlighting the need for healthcare system-specific approaches to risk mitigation. Given the observed differences in patient populations, institutional characteristics, magnitude of risk, and subgroup analyses, we recommend developing tailored intervention strategies that account for each system’s unique contextual factors, resources, and constraints while maintaining vigilance regarding opioid-associated mortality risk.

After 30 days, the mortality risk associated with opioid initiation began to stabilize across settings. While previous studies have demonstrated elevated risks of opioid-related adverse effects during the initial phase of treatment in elderly populations,^11,13,22^ these comparisons were between opioid-exposed and unexposed patients and lacked a specific timeframe. Our study took a novel approach by comparing new users to consistent users within the exposed group, identifying a specific 30-day window of heightened risk, providing clinicians with an actionable timeframe for intervention. This 30-day period suggests a timeframe for close monitoring of patients newly initiating opioid therapy, potentially through weekly follow-up for adverse events and increased patient and caregiver education about monitoring opioid-related adverse effects.

Pneumonia emerged as a significant factor in the association between short-term mortality and opioid initiation. In the primary cohort, we found a particularly high prevalence of pneumonia among new users as a potential cause of early mortality, based on GPT-extracted causes of death. This prompted us to investigate whether the association reflected pneumonia resulting from opioid exposure, as suggested by previous studies,^12,22,23^ or preexisting conditions. Our supplementary analyses revealed contrasting patterns between cohorts. In the primary cohort, patients with preexisting pneumonia showed a 2·38-fold increase in short-term mortality after opioid initiation, while opioid initiation did not increase subsequent pneumonia risk among those without preexisting pneumonia. The validation cohort showed an opposite pattern: opioid initiation was associated with 1·2 fold increased risk of subsequent pneumonia, but not with increased mortality among patients with preexisting pneumonia. These contrasting findings likely reflect the interaction between opioid-induced respiratory depression and pneumonia manifesting differently across healthcare settings - exacerbating existing pneumonia in the primary cohort while increasing susceptibility to new infections in the validation cohort.^10,24,25^ These differences highlight the need for site-specific risk mitigation strategies. Our analysis did not examine specific opioid dose-response relationships in relation to adverse events, which may explain the disparate findings across healthcare settings. Nonetheless, our findings emphasize the importance of monitoring for pneumonia among patients with dementia who are newly prescribed opioids, and also carefully considering the risks and benefits of opioid prescribing among opioid-naive patients with co-morbid dementia and pneumonia.

The integration of LLM-based clinical note analysis with traditional structured data analysis strengthened our research methodology. While structured data analysis established the mortality patterns, LLM analysis of clinical notes identified pneumonia as a significant factor, enabling targeted supplementary analyses that revealed complex relationships between pneumonia timing, opioid initiation, and mortality across healthcare settings. This complementary approach demonstrates how unstructured clinical notes can provide crucial context often missed in structured data alone, allowing us to generate and test hypothesis about complex clinical outcomes.

Our study has several important limitations. First, while we obtained death dates from both EHR and state death records, we could not access clinical notes from outside facilities, potentially limiting our understanding of health status at death for patients who died in other healthcare settings. Second, we could only verify medication administration for inpatient care: our outpatient data only captured prescriptions and dispensing records, and not actual adherence. Third, our use of GPT for analyzing clinical notes was limited to the primary cohort. Fourth, the higher mortality risk among new users could be partially attributed to acute severe conditions necessitating first-time opioid use, such as traumatic injuries or accidents. However, our GPT analysis of clinical notes from the primary cohort revealed that such acute traumatic events were relatively uncommon causes of death, suggesting that this potential confounding factor may not fully explain the observed mortality difference.

## 5. Conclusion

This study demonstrates significantly elevated short-term mortality risk associated with initiating opioid use in patients with dementia, particularly within the first 14 days and stabilizing by day 30. The consistency of these findings across distinctly different healthcare settings, despite varying magnitude of risks, strengthens their generalizability while highlighting the need for setting-specific risk mitigation strategies. The heightened vulnerability of opioid-naïve patients to adverse effects emphasizes the importance of careful risk assessment and close monitoring when initiating opioid therapy in this vulnerable population.

## Contributors

Yeon-Mi Hwang: Conceptualization, Resources, Data Curation, Software, Formal Analysis, Investigation, Visualization, Data Interpretation, Methodology, Writing - original draft, Writing - review and editing; Jennifer M. Hah: Conceptualization, Methodology, Data Interpretation, Writing - review and editing; Jennifer E. Bramen: Methodology, Data Interpretation, Writing - review and editing; Jennifer J. Hadlock: Methodology, Data Interpretation, Writing - review and editing, Administrative or technical support; Tina Hernandez-Boussard: Conceptualization, Supervision, Writing - review and editing, Administrative, technical, or material support; YH and TH-B had full access to the primary cohort data in the study and verified the data. YH and JJH had full access to the validation cohort data in the study and verified the data.

## Data Sharing

Data cannot be shared for ethical/privacy reasons. However, all clinical logic has been transparently shared. The results have been aggregated and reported in this manuscript to the fullest extent possible while adhering to legal requirements for protecting personal health information. The code is available in the GitHub repository: https://github.com/su-boussard-lab/dementia-opioid-initiation/tree/main

## Data Availability

All clinical logic has been shared. Results have been aggregated and reported within this paper to the extent possible while maintaining privacy from personal health information as required by law.

## Acknowledgements

During the preparation of this work, the author used Claude 3.5 Sonnet, to assist with improving text readability. All AI-generated suggestions were carefully evaluated, and only selected revisions that preserved the scientific integrity and intended meaning were incorporated. The author critically reviewed each modification, performed additional editing to refine the content further, and maintained full editorial control and responsibility for the final content. The following prompt was used: “I have a paragraph of a scientific manuscript that I would like help improving the writing and readability of. I will paste the text below. Please review it and suggest edits to make the writing clearer, more concise, and easier to follow, while preserving the scientific content and meaning. Focus on improving awkward phrasing, run-on sentences, unnecessary jargon, grammar and punctuation. Do not alter the core scientific ideas or add any new information. [Paste manuscript text]. Please provide a brief explanation for your changes.”

