## Supplementary materials for "Short-Term Mortality After Opioid Initiation Among Opioid-Naïve and Non-Naïve Patients with Dementia: A Retrospective Cohort Study"

**Table S1:** ICD-10 codes and search criteria of dementia and its subcategories

**Table S2:** RxNorm codes of opioid ingredient

**Table S3:** Variable definition

**Table S4:** ICD-10 codes and inclusion criteria of comorbidities

**Table S5:** RxNorm codes, categories, and inclusion criteria of medications

**Table S6:** Secure GPT-3.5 Turbo prompt for potential causes of death identification

**Table S7:** Descriptive statistics of opioid-exposed patients, comparing new users and consistent users in primary cohort on comorbidities and medication exposure

**Table S8:** Descriptive statistics of the dementia/mild cognitive impairment cohort based on post-diagnosis opioid exposure status

**Table S9:** Descriptive statistics of opioid-exposed patients, comparing new users and consistent users in validation cohort

**Table S10**: Notes, GPT Responses, and Health Condition Categories of Mismatches Between Specified Principal Diagnoses at the Time of Death and GPT Responses

**Figure S1:** Flowchart of cohort and exposure group selection in the validation cohort

**Figure S2:** 180-day survival probability curves comparing new users and consistent users in primary and validation cohorts

**Figure S3:** Sensitivity analysis comparing first-time opioid users to long-term consistent users in primary and validation cohort

**Figure S4:** Strong opioid-specific and weak-opioid specific 14-day survival probability curve comparing new users and consistent users in primary and validation cohort

**Figure S5:** MCI-specific and dementia-specific 14-day survival probability curve comparing new users and consistents users in primary and validation cohort

**Figure S6**: Inpatient-specific and outpatient-specific 14-day survival probability curve comparing new users and consistents users in primary and validation cohort

**Figure S7**: Post-hoc supplementary analysis of short-term mortality associated with opioid initiation in patients with preexising pneumonia and pneumonia development risk following opioid initiation in patients without preexisting pneumonia in primary and validation cohort

| **Diagnosis** | **ICD-10^a^** | **Diagnosis name search** |
| --- | --- | --- |
| **Mild cognitive impairment** | G31.84 | “mild cognitive impairment” |
| **Dementia** | F00, F01, F02, F03, G30, F05.1, G31.0, G31.83, G31.1 | “dementia” |
| **Alzheimer’s diseases** | G30 | “alzheimer” |
| **Frontotemporal dementia** | G31.0 | “frontotemporal” |
| **Vascular dementia** | F01 | “vascular dementia” |
| **Lewy body dementia** | G31.83 | “lewy body” |

**Table S1** ICD-10 codes and search criteria of dementia and its subcategories

^a^Descendant codes of the listed ICD-10 codes are included

| **Opioid ingredient** | **RxNorm** |
| --- | --- |
| **codeine** | 2670 |
| **hydrocodone** | 5489 |
| **fentanyl** | 4337 |
| **hydromorphone** | 3423 |
| **oxycodone** | 7804 |
| **morphine** | 7052 |
| **methadone** | 6813 |
| **tramadol** | 10689 |
| **meperidine** | 6754 |

**Table S2** RxNorm codes of opioid ingredient

| **Category** | **Variable** | **Definitions** |
| --- | --- | --- |
| **Patient Characteristics** | Sex | Sex noted in medical record; Female, Male, Other |
|  | Age At Diagnosis | Age at the first dementia or mild cognitive impairment diagnosis. |
|  | Race | Self-reported race noted in medical record; White, Black, Asian, Native American, Hawaiian/Pacific Islander, Other, Unknown/Refused. |
|  | Ethnicity | Self-reported ethnicity noted in medical record; Hispanic/Latino, Non-Hispanic/Non-Latino, Unknown/Refused |
|  | Bmi | The most recent Body Mass Index (BMI, kg/m²) recorded before the patient's first opioid exposure was used. Extreme values exceeding world record thresholds were considered invalid and imputed as missing. For analysis, BMI was categorized as an ordinal variable with the following classifications: Underweight (<18.5) = 1, Normal (18.5–24.9) = 2, Overweight (25.0–29.9) = 3, Obese (30.0–34.9) = 4, and Severely Obese (>35.0) = 5. |
|  | Insurance | Type of insurance effective at the time of the first opioid exposure; Medicaid/Medicare, Private, Other |
| **Death** | Death | Patient’s death status as of the end of the study period (July 31, 2024), recorded as a binary variable {0 = alive, 1 = deceased}. |
|  | Death Within 14 Days After First Opioid Exposure | Patient’s death status on follow-up day 14, recorded as a binary variable {0 = alive, 1 = deceased}. |
|  | Death Within 60 Days After First Opioid Exposure | Patient’s death status on follow-up day 60, recorded as a binary variable {0 = alive, 1 = deceased}. |
|  | Death Within 180 Days After First Opioid Exposure | Patient’s death status on follow-up day 180, recorded as a binary variable {0 = alive, 1 = deceased}. |
|  | Days To Death From First Opioid Exposure | Number of days from the first opioid exposure to the date of death |
| **Other** | Last Encounter Date | Date of the patient’s last recorded encounter. For deceased patients, the date of death is considered the last encounter date. |

**Table S3** Variable definition

| **Diagnosis** | **ICD-10**^a^ | **Inclusion Criteria**^b^ |
| --- | --- | --- |
| **myocardial infarction** | I21, I22, I25.2 | mortality risk factor |
| **congestive heart failure** | I09.9, I11.0, I13.0, I13.2, I25.5, I42.0, I42.5, I42.6, I42.7, I42.8, I42.9, I43, I50, P29.0 | mortality risk factor |
| **peripheral vascular disease** | I70, I71, I73.1, I73.8, I73.9, I77.1, I79.0, I79.2, K55.1, K55.8, K55.9, Z95.8, Z95.9 | mortality risk factor |
| **cerebrovascular disease** | G45, G46, I60, I61, I62, I63, I64, I65, I66, I67, I68, I69, H34.0 | mortality risk factor |
| **chronic pulmonary disease** | I27.8, I27.9, J40, J41, J42, J43, J44, J45, J46, J47, J60, J61, J62, J63, J64, J65, J66, J67, J68.4, J70.1, J70.3 | mortality risk factor |
| **rheumatologic disease** | M05, M06, M31.5, M32, M33, M34, M35.1, M35.3, M36.0 | mortality risk factor |
| **peptic ulcer** | K25, K26, K27, K28 | mortality risk factor |
| **hemiplegia/paraplegia** | G04.1, G11.4, G80.1, G80.2, G81, G82, G83.0, G83.1, G83.2, G83.3, G83.4, G83.9 | mortality risk factor |
| **diabetes without complications** | E10.0, E10.1, E10.6, E10.8, E10.9, E11.0, E11.1, E11.6, E11.8, E11.9, E12.0, E12.1, E12.6, E12.8, E12.9, E13.0, E13.1, E13.6, E13.8, E13.9, E14.0, E14.1, E14.6, E14.8, E14.9 | mortality risk factor |
| **diabetes with chronic complications** | E10.2, E10.3, E10.4, E10.5, E10.7, E11.2, E11.5, E11.7, E12.2, E12.3, E12.4, E12.5, E12.7, E13.2, E13.3, E13.4, E13.5, E13.7, E14.2, E14.3, E14.4, E14.5, E14.7 | mortality risk factor |
| **mild liver diseases** | B18, K70.0, K70.1, K70.2, K70.3, K70.9, K71.3, K71.4, K71.5, K71.7, K73, K76.0, K76.2, K76.3, K76.4, K76.8, K76.9, Z94.4 | mortality risk factor |
| **moderate/severe liver disease** | I85.0, I85.9, I86.4, I98.2, K70.4, K71.1, K72.1, K72.9, K76.5, K76.6, K76.7 | mortality risk factor |
| **renal disease** | I20, I131, N03.2, N03.3, N03.4, N03.5, N03.6, N03.7, N05.2, N05.3, N05.4, N05.5, N05.6, N05.7, N18, N19, N25.0, Z49.0, Z49.1, Z49.2, Z94.0, N99.2 | mortality risk factor |
| **any malignancy (tumor, leukemia, lymphoma)** | C00, C01, C02, C03, C04, C05, C06, C07, C08, C09, C10, C11, C12, C13, C14, C15, C16, C17, C18, C19, C20, C21, C22, C23, C24, C25, C26, C30, C31, C32, C33, C34, C37, C38, C39, C40, C41, C43, C45, C46, C47, C48, C49, C50, C51, C52, C53, C54, C55, C56, C57, C58, C60, C61, C62, C63, C64, C65, C66, C67, C68, C69, C70, C71, C72, C73, C74, C75, C76, C81, C82, C83, C84, C85, C88, C90, C91, C92, C93, C94, C95, C96, C97 | mortality risk factor |
| **metastatic solid tumor** | C77, C78, C79, C80 | mortality risk factor |
| **HIV/AIDS** | B20, B21, B22, B24 | mortality risk factor |
| **cognitive decline** | R41.81, R41.89 | dementia risk factor |
| **hypertension** | I10, I11, I12, I13, I15, I16 | dementia risk factor |
| **hyperlipidemia** | E78.2, E78.4, E78.5 | dementia risk factor |
| **excessive alcohol** | F10 | dementia risk factor |
| **atherosclerosis** | I25.1, I70 | dementia risk factor |
| **hypercholesterolemia** | E78.0 | dementia risk factor |
| **atrial fibrillation** | I48 | dementia risk factor |
| **traumatic brain injury** | S02.0, S02.1, S02.8, S02.91, S04.02, S04.03, S04.04, S06, S07.1, T74.4 | dementia risk factor |
| **hearing loss** | H90, H91 | dementia risk factor |
| **sleep apnea** | G47.3 | dementia risk factor |
| **prediabetes** | R73.03 | dementia risk factor |
| **delirium** | R41.0 | dementia risk factor |
| **depression** | F32.0, F32.1, F32.2, F32.3, F32.4, F32.5, F32.89, F32.9, F33.1, F33.3, F33.4, F33.9 | dementia risk factor |
| **schizophrenia/mood psychotic disorder** | F20, F21, F22, F23, F24, F25, F27, F28, F29 | dementia risk factor |
| **mood disorder** | F30, F31, F32, F33, F34, F39 | dementia risk factor |
| **anxiety and non-psychotic mental disorder** | F40, F41, F42, F43, F44, F45, F48 | dementia risk factor |
| **behavioral syndromes associated with physiological disturbances and physical factors** | F50, F51, F52, F53, F54, F55, F59 | dementia risk factor |
| **disorders of adult personality and behavior** | F60, F63, F64, F65, F66, F68, F69 | dementia risk factor |
| **intellectual disability** | F70, F71, F72, F73, F78, F79 | dementia risk factor |
| **developmental disorder** | F80, F81, F82, F84, F88, F89 | dementia risk factor |
| **behavioral and emotional disorder with onset in early ages** | F90, F91, F93, F94, F95, F98 | dementia risk factor |

**Table S4**. ICD-10 codes and inclusion criteria of comorbidities.

^a^Descendant codes of the listed ICD-10 codes are included

^b^Mortality risk factors were selected based on Charlson’s comorbidity index. Dementia risk factors were selected based on a literature review.

| **Medication ingredient** | **RxNorm Code** | **Therapeutic/chemical subcategory** | **Therapeutic category** | **Inclusion criteria** |
| --- | --- | --- | --- | --- |
| donepezil | 135447 | **acetylcholinesterase inhibitor** | antidementia medication | antidementia medication |
| rivastigmine | 183379 | **acetylcholinesterase inhibitor** | antidementia medication | antidementia medication |
| galantamine | 4637 | **acetylcholinesterase inhibitor** | antidementia medication | antidementia medication |
| tacrine | 10318 | **acetylcholinesterase inhibitor** | antidementia medication | antidementia medication |
| memantine | 6719 | **acetylcholinesterase inhibitor** | antidementia medication | antidementia medication |
| donepezil/memantine | 1430990 | **acetylcholinesterase inhibitor** | antidementia medication | antidementia medication |
| brompheniramine | 1767 | **first generation antihistamine** | antihistamine | AGS Beers Criteria |
| chlorpheniramine | 2400 | **first generation antihistamine** | antihistamine | AGS Beers Criteria |
| cyproheptadine | 3013 | **first generation antihistamine** | antihistamine | AGS Beers Criteria |
| dimenhydrinate | 3444 | **first generation antihistamine** | antihistamine | AGS Beers Criteria |
| diphenhydramine | 3498 | **first generation antihistamine** | antihistamine | AGS Beers Criteria |
| doxylamine | 3642 | **first generation antihistamine** | antihistamine | AGS Beers Criteria |
| hydroxyzine | 5553 | **first generation antihistamine** | antihistamine | AGS Beers Criteria |
| meclizine | 6676 | **first generation antihistamine** | antihistamine | AGS Beers Criteria |
| promethazine | 8745 | **first generation antihistamine** | antihistamine | AGS Beers Criteria |
| triprolidine | 10849 | **first generation antihistamine** | antihistamine | AGS Beers Criteria |
| nitrofurantoin | 7454 | **nitrofurantoin** | anti-infective | AGS Beers Criteria |
| aspirin | 1191 | **aspirin** | cardiovascular and antithrombotic | AGS Beers Criteria |
| warfarin | 11289 | **warfarin** | cardiovascular and antithrombotic | AGS Beers Criteria |
| rivaroxaban | 1114195 | **rivaroxaban** | cardiovascular and antithrombotic | AGS Beers Criteria |
| dipyridamole | 3521 | **dipyridamole** | cardiovascular and antithrombotic | AGS Beers Criteria |
| doxazosin | 49276 | **non-selective peripheral alpha-1 blockers** | cardiovascular and antithrombotic | AGS Beers Criteria |
| prazosin | 8629 | **non-selective peripheral alpha-1 blockers** | cardiovascular and antithrombotic | AGS Beers Criteria |
| terazosin | 37798 | **non-selective peripheral alpha-1 blockers** | cardiovascular and antithrombotic | AGS Beers Criteria |
| clonidine | 2599 | **central alpha-agonist** | cardiovascular and antithrombotic | AGS Beers Criteria |
| guanfacine | 40114 | **central alpha-agonist** | cardiovascular and antithrombotic | AGS Beers Criteria |
| nifedipine | 7417 | **nifedipine** | cardiovascular and antithrombotic | AGS Beers Criteria |
| amiodarone | 703 | **amiodarone** | cardiovascular and antithrombotic | AGS Beers Criteria |
| digoxin | 3407 | **digoxin** | cardiovascular and antithrombotic | AGS Beers Criteria |
| amitriptyline | 704 | **TCAs** | central nervous system | AGS Beers Criteria |
| amoxapine | 722 | **TCAs** | central nervous system | AGS Beers Criteria |
| clomipramine | 2597 | **TCAs** | central nervous system | AGS Beers Criteria |
| desipramine | 3247 | **TCAs** | central nervous system | AGS Beers Criteria |
| doxepin | 3638 | **TCAs** | central nervous system | AGS Beers Criteria |
| imipramine | 5691 | **TCAs** | central nervous system | AGS Beers Criteria |
| nortriptyline | 7531 | **TCAs** | central nervous system | AGS Beers Criteria |
| paroxetine | 32937 | **SSRIs** | central nervous system | AGS Beers Criteria |
| benztropine | 1424 | **antiparkinsoniac agent** | central nervous system | AGS Beers Criteria |
| trihexyphenidyl | 10811 | **antiparkinsoniac agent** | central nervous system | AGS Beers Criteria |
| aripiprazole | 89013 | **antipsychotics** | central nervous system | AGS Beers Criteria |
| haloperidol | 5093 | **antipsychotics** | central nervous system | AGS Beers Criteria |
| olanzapine | 61381 | **antipsychotics** | central nervous system | AGS Beers Criteria |
| quetiapine | 51272 | **antipsychotics** | central nervous system | AGS Beers Criteria |
| risperidone | 35636 | **antipsychotics** | central nervous system | AGS Beers Criteria |
| butalbital | 19860 | **barbiturates** | central nervous system | AGS Beers Criteria |
| phenobarbital | 8134 | **barbiturates** | central nervous system | AGS Beers Criteria |
| primidone | 8691 | **barbiturates** | central nervous system | AGS Beers Criteria |
| alprazolam | 596 | **benzodiazepines** | central nervous system | AGS Beers Criteria |
| chlordiazepoxide | 2356 | **benzodiazepines** | central nervous system | AGS Beers Criteria |
| clobazam | 21241 | **benzodiazepines** | central nervous system | AGS Beers Criteria |
| clonazepam | 2598 | **benzodiazepines** | central nervous system | AGS Beers Criteria |
| clorazepate | 2353 | **benzodiazepines** | central nervous system | AGS Beers Criteria |
| diazepam | 3322 | **benzodiazepines** | central nervous system | AGS Beers Criteria |
| estazolam | 4077 | **benzodiazepines** | central nervous system | AGS Beers Criteria |
| lorazepam | 6470 | **benzodiazepines** | central nervous system | AGS Beers Criteria |
| midazolam | 6960 | **benzodiazepines** | central nervous system | AGS Beers Criteria |
| oxazepam | 7781 | **benzodiazepines** | central nervous system | AGS Beers Criteria |
| temazepam | 10355 | **benzodiazepines** | central nervous system | AGS Beers Criteria |
| triazolam | 10767 | **benzodiazepines** | central nervous system | AGS Beers Criteria |
| eszopiclone | 461016 | **nonbenzodiazepine benzodiazepine receptor agonist hypnotics** | central nervous system | AGS Beers Criteria |
| zaleplon | 74667 | **nonbenzodiazepine benzodiazepine receptor agonist hypnotics** | central nervous system | AGS Beers Criteria |
| zolpidem | 39993 | **nonbenzodiazepine benzodiazepine receptor agonist hypnotics** | central nervous system | AGS Beers Criteria |
| meprobamate | 6760 | **meprobamate** | central nervous system | AGS Beers Criteria |
| ergoid mesylates | 4024 | **ergoid mesylates** | central nervous system | AGS Beers Criteria |
| methyltestosterone | 6904 | **androgen** | endocrine | AGS Beers Criteria |
| testosterone | 10379 | **androgen** | endocrine | AGS Beers Criteria |
| estrogens | 4100 | **estrogens** | endocrine | AGS Beers Criteria |
| bazedoxifene | 1441386 | **estrogens** | endocrine | AGS Beers Criteria |
| bazedoxifene / estrogens, conjugated (USP) | 1441391 | **estrogens** | endocrine | AGS Beers Criteria |
| dienestrol | 3368 | **estrogens** | endocrine | AGS Beers Criteria |
| dienogest | 22968 | **estrogens** | endocrine | AGS Beers Criteria |
| dienogest / estradiol | 994203 | **estrogens** | endocrine | AGS Beers Criteria |
| estradiol | 4083 | **estrogens** | endocrine | AGS Beers Criteria |
| estradiol / norgestimate | 261414 | **estrogens** | endocrine | AGS Beers Criteria |
| estradiol cypionate | 1000146 | **estrogens** | endocrine | AGS Beers Criteria |
| estradiol valerate | 24395 | **estrogens** | endocrine | AGS Beers Criteria |
| estrogens, conjugated (USP) | 4099 | **estrogens** | endocrine | AGS Beers Criteria |
| estrogens, conjugated (USP) / medroxyprogesterone | 1006917 | **estrogens** | endocrine | AGS Beers Criteria |
| estrogens, esterified (USP) | 214549 | **estrogens** | endocrine | AGS Beers Criteria |
| estropipate | 33747 | **estrogens** | endocrine | AGS Beers Criteria |
| inert ingredients | 748794 | **estrogens** | endocrine | AGS Beers Criteria |
| medroxyprogesterone | 6691 | **estrogens** | endocrine | AGS Beers Criteria |
| medroxyprogesterone acetate | 1000112 | **estrogens** | endocrine | AGS Beers Criteria |
| norgestimate | 31994 | **estrogens** | endocrine | AGS Beers Criteria |
| insulin isophane, human | 253181 | **insulin** | endocrine | AGS Beers Criteria |
| insulin aspart protamine, human / insulin aspart, human | 1007184 | **insulin** | endocrine | AGS Beers Criteria |
| insulin isophane / insulin, regular, human | 1008501 | **insulin** | endocrine | AGS Beers Criteria |
| insulin lispro protamine, human | 816726 | **insulin** | endocrine | AGS Beers Criteria |
| insulin isophane | 1605101 | **insulin** | endocrine | AGS Beers Criteria |
| insulin, regular, human | 253182 | **insulin** | endocrine | AGS Beers Criteria |
| insulin lispro protamine, human | 314684 | **insulin** | endocrine | AGS Beers Criteria |
| insulin aspart protamine, human | 352385 | **insulin** | endocrine | AGS Beers Criteria |
| insulin aspart, human | 51428 | **insulin** | endocrine | AGS Beers Criteria |
| insulin lispro | 86009 | **insulin** | endocrine | AGS Beers Criteria |
| gliclazide | 4816 | **sulfonylureas** | endocrine | AGS Beers Criteria |
| glimepiride | 25789 | **sulfonylureas** | endocrine | AGS Beers Criteria |
| glipizide | 4821 | **sulfonylureas** | endocrine | AGS Beers Criteria |
| glyburide | 4815 | **sulfonylureas** | endocrine | AGS Beers Criteria |
| desiccated thyroid | 10572 | **thyroid** | endocrine | AGS Beers Criteria |
| megestrol | 6703 | **megestrol** | endocrine | AGS Beers Criteria |
| somatropin | 61148 | **growth hormone** | endocrine | AGS Beers Criteria |
| lonapegsomatropin | 2569562 | **growth hormone** | endocrine | AGS Beers Criteria |
| dexlansoprazole | 816346 | **proton-pump inhibitors** | gastrointestinal | AGS Beers Criteria |
| esomeprazole | 283742 | **proton-pump inhibitors** | gastrointestinal | AGS Beers Criteria |
| lansoprazole | 17128 | **proton-pump inhibitors** | gastrointestinal | AGS Beers Criteria |
| omeprazole | 7646 | **proton-pump inhibitors** | gastrointestinal | AGS Beers Criteria |
| pantoprazole | 40790 | **proton-pump inhibitors** | gastrointestinal | AGS Beers Criteria |
| rabeprazole | 114979 | **proton-pump inhibitors** | gastrointestinal | AGS Beers Criteria |
| metoclopramide | 6915 | **metoclopramide** | gastrointestinal | AGS Beers Criteria |
| atropine | 1223 | **GI antispasmodics** | gastrointestinal | AGS Beers Criteria |
| clidinium/chlordiazepoxide | 611854 | **GI antispasmodics** | gastrointestinal | AGS Beers Criteria |
| dicyclomine | 3361 | **GI antispasmodics** | gastrointestinal | AGS Beers Criteria |
| hyoscyamine | 153970 | **GI antispasmodics** | gastrointestinal | AGS Beers Criteria |
| scopolamine | 9601 | **GI antispasmodics** | gastrointestinal | AGS Beers Criteria |
| mineral oil | 6972 | **mineral oil** | gastrointestinal | AGS Beers Criteria |
| desmopressin | 3251 | **desmopressin** | genitourinary | AGS Beers Criteria |
| aspirin | 1191 | **non-COX-2 selective NSAIDS** | pain medications | AGS Beers Criteria |
| diclofenac | 3355 | **non-COX-2 selective NSAIDS** | pain medications | AGS Beers Criteria |
| diflunisal | 3393 | **non-COX-2 selective NSAIDS** | pain medications | AGS Beers Criteria |
| etodolac | 24605 | **non-COX-2 selective NSAIDS** | pain medications | AGS Beers Criteria |
| flurbiprofen | 4502 | **non-COX-2 selective NSAIDS** | pain medications | AGS Beers Criteria |
| ibuprofen | 5640 | **non-COX-2 selective NSAIDS** | pain medications | AGS Beers Criteria |
| indomethacin | 5781 | **non-COX-2 selective NSAIDS** | pain medications | AGS Beers Criteria |
| ketorolac | 35827 | **non-COX-2 selective NSAIDS** | pain medications | AGS Beers Criteria |
| meloxicam | 41493 | **non-COX-2 selective NSAIDS** | pain medications | AGS Beers Criteria |
| nabumetone | 31448 | **non-COX-2 selective NSAIDS** | pain medications | AGS Beers Criteria |
| naproxen | 7258 | **non-COX-2 selective NSAIDS** | pain medications | AGS Beers Criteria |
| oxaprozin | 32613 | **non-COX-2 selective NSAIDS** | pain medications | AGS Beers Criteria |
| piroxicam | 8356 | **non-COX-2 selective NSAIDS** | pain medications | AGS Beers Criteria |
| sulindac | 10237 | **non-COX-2 selective NSAIDS** | pain medications | AGS Beers Criteria |
| carisoprodol | 2101 | **skeletal muscle relaxant** | pain medications | AGS Beers Criteria |
| chlorzoxazone | 2410 | **skeletal muscle relaxant** | pain medications | AGS Beers Criteria |
| cyclobenzaprine | 21949 | **skeletal muscle relaxant** | pain medications | AGS Beers Criteria |
| metaxalone | 59078 | **skeletal muscle relaxant** | pain medications | AGS Beers Criteria |
| methocarbamol | 6845 | **skeletal muscle relaxant** | pain medications | AGS Beers Criteria |
| orphenadrine | 7715 | **skeletal muscle relaxant** | pain medications | AGS Beers Criteria |
| dabigatran | 1546356 | **blood thinner** | blood thinner | AGS Beers PIP |
| prasugrel | 613391 | **blood thinner** | blood thinner | AGS Beers PIP |
| ticagrelor | 1116632 | **blood thinner** | blood thinner | AGS Beers PIP |
| mirtazapine | 15996 | **TeCA** | antidepressants | AGS Beers PIP |
| desvenlafaxine | 734064 | **SNRIs** | antidepressants | AGS Beers PIP |
| duloxetine | 72625 | **SNRIs** | antidepressants | AGS Beers PIP |
| levomilnacipran | 1433212 | **SNRIs** | antidepressants | AGS Beers PIP |
| milnacipran | 588250 | **SNRIs** | antidepressants | AGS Beers PIP |
| venlafaxine | 39786 | **SNRIs** | antidepressants | AGS Beers PIP |
| citalopram | 2556 | **SSRIs** | antidepressants | AGS Beers PIP |
| escitaloram | 321988 | **SSRIs** | antidepressants | AGS Beers PIP |
| fluoxetine | 4493 | **SSRIs** | antidepressants | AGS Beers PIP |
| fluvoxamine | 42355 | **SSRIs** | antidepressants | AGS Beers PIP |
| nefazodone | 31565 | **SSRIs** | antidepressants | AGS Beers PIP |
| paroxetine | 32937 | **SSRIs** | antidepressants | AGS Beers PIP |
| sertraline | 36437 | **SSRIs** | antidepressants | AGS Beers PIP |
| trazodone | 10737 | **SSRIs** | antidepressants | AGS Beers PIP |
| amitriptyline | 704 | **TCAs** | antidepressants | AGS Beers PIP |
| amoxapine | 722 | **TCAs** | antidepressants | AGS Beers PIP |
| clomipramine | 2597 | **TCAs** | antidepressants | AGS Beers PIP |
| desipramine | 3247 | **TCAs** | antidepressants | AGS Beers PIP |
| doxepine | 3638 | **TCAs** | antidepressants | AGS Beers PIP |
| imipramine | 5691 | **TCAs** | antidepressants | AGS Beers PIP |
| nortriptyline | 7531 | **TCAs** | antidepressants | AGS Beers PIP |
| protriptyline | 8886 | **TCAs** | antidepressants | AGS Beers PIP |
| trimipramine | 10834 | **TCAs** | antidepressants | AGS Beers PIP |
| carbamazepine | 2002 | **anticonvulsants** | antiepileptics | AGS Beers PIP |
| oxcarbazepine | 32624 | **anticonvulsants** | antiepileptics | AGS Beers PIP |
| aripiprazole | 89013 | **antipsychotics** | antipsychotics | AGS Beers PIP |
| aripiprazole lauroxil | 1673265 | **antipsychotics** | antipsychotics | AGS Beers PIP |
| asenapine | 784649 | **antipsychotics** | antipsychotics | AGS Beers PIP |
| brexpiprazole | 1658314 | **antipsychotics** | antipsychotics | AGS Beers PIP |
| cariprazine | 1667655 | **antipsychotics** | antipsychotics | AGS Beers PIP |
| clozapine | 2626 | **antipsychotics** | antipsychotics | AGS Beers PIP |
| haloperidol | 5093 | **antipsychotics** | antipsychotics | AGS Beers PIP |
| iloperidone | 73178 | **antipsychotics** | antipsychotics | AGS Beers PIP |
| lurasidone | 1040028 | **antipsychotics** | antipsychotics | AGS Beers PIP |
| molindone | 7019 | **antipsychotics** | antipsychotics | AGS Beers PIP |
| olanzapine | 61381 | **antipsychotics** | antipsychotics | AGS Beers PIP |
| paliperidone | 679314 | **antipsychotics** | antipsychotics | AGS Beers PIP |
| pimavanserin | 1791685 | **antipsychotics** | antipsychotics | AGS Beers PIP |
| pimozide | 8331 | **antipsychotics** | antipsychotics | AGS Beers PIP |
| quetiapine | 51272 | **antipsychotics** | antipsychotics | AGS Beers PIP |
| risperidone | 35636 | **antipsychotics** | antipsychotics | AGS Beers PIP |
| thiothixene | 10510 | **antipsychotics** | antipsychotics | AGS Beers PIP |
| ziprasidone | 115698 | **antipsychotics** | antipsychotics | AGS Beers PIP |
| amiloride | 644 | **diuretics** | diuretics | AGS Beers PIP |
| amiloride_hydrochlorothiazide | 214212 | **diuretics** | diuretics | AGS Beers PIP |
| amiloride hydrochloride | 142424 | **diuretics** | diuretics | AGS Beers PIP |
| bumetanide | 1808 | **diuretics** | diuretics | AGS Beers PIP |
| chlorothiazide | 2396 | **diuretics** | diuretics | AGS Beers PIP |
| chlorthalidone | 2409 | **diuretics** | diuretics | AGS Beers PIP |
| conivaptan | 302285 | **diuretics** | diuretics | AGS Beers PIP |
| conivaptan hydrochloride | 1294548 | **diuretics** | diuretics | AGS Beers PIP |
| eplerenone | 298869 | **diuretics** | diuretics | AGS Beers PIP |
| ethacrynate | 62349 | **diuretics** | diuretics | AGS Beers PIP |
| ethacrynate sodium | 4108 | **diuretics** | diuretics | AGS Beers PIP |
| ethacrynic acid | 4109 | **diuretics** | diuretics | AGS Beers PIP |
| finerenone | 2562811 | **diuretics** | diuretics | AGS Beers PIP |
| furosemide | 4603 | **diuretics** | diuretics | AGS Beers PIP |
| hydrochlorothiazide | 5487 | **diuretics** | diuretics | AGS Beers PIP |
| hydrochlorothiazide/spironolactone | 324042 | **diuretics** | diuretics | AGS Beers PIP |
| hydrochlorothiazide/triamterene | 258337 | **diuretics** | diuretics | AGS Beers PIP |
| indapamide | 5764 | **diuretics** | diuretics | AGS Beers PIP |
| methyclothiazide | 6860 | **diuretics** | diuretics | AGS Beers PIP |
| metolazone | 6916 | **diuretics** | diuretics | AGS Beers PIP |
| spironolactone | 9997 | **diuretics** | diuretics | AGS Beers PIP |
| tolvaptan | 358257 | **diuretics** | diuretics | AGS Beers PIP |
| torsemide | 38413 | **diuretics** | diuretics | AGS Beers PIP |
| triamterene | 10763 | **diuretics** | diuretics | AGS Beers PIP |
| dextromethorphan/quinidine | 1040053 | **dextromethorphan/quinidine** | dextromethorphan/quinidine | AGS Beers PIP |
| sulfamethoxazole/trimethoprim | 10831 | **sulfamethoxazole/trimethoprim** | sulfamethoxazole/trimethoprim | AGS Beers PIP |
| canigliflozin | 1373458 | **SGLT2** | SGLT2 | AGS Beers PIP |
| dapagliflozin | 1488564 | **SGLT2** | SGLT2 | AGS Beers PIP |
| emplaglifozin | 1545653 | **SGLT2** | SGLT2 | AGS Beers PIP |
| ertuglifozin | 1992672 | **SGLT2** | SGLT2 | AGS Beers PIP |
| acetaminophen | 161 | **acetaminophen** | pain medications | Clinician-recommended relevance |
| gabapentin | 25480 | **anticonvulsants** | antiepileptics | Clinician-recommended relevance |
| pregabalin | 187832 | **anticonvulsants** | antiepileptics | Clinician-recommended relevance |
| topiramate | 38404 | **anticonvulsants** | antiepileptics | Clinician-recommended relevance |
| lamotrigrine | 28439 | **phenyltriazine** | antiepileptics | Clinician-recommended relevance |
| levetiracetam | 114477 | **anticonvulsants** | antiepileptics | Clinician-recommended relevance |
| phenytoin | 8183 | **anticonvulsants** | antiepileptics | Clinician-recommended relevance |
| valproic acid | 11118 | **anticonvulsants** | antiepileptics | Clinician-recommended relevance |
| zonisamide | 39998 | **anticonvulsants** | antiepileptics | Clinician-recommended relevance |
| tiagabine | 31914 | **anticonvulsants** | antiepileptics | Clinician-recommended relevance |
| verapamil | 11170 | **calcium channel blockers** | antihypertensives | Clinician-recommended relevance |
| sumatriptan | 37418 | **triptan** | antimigraine | Clinician-recommended relevance |
| naratriptan | 141366 | **triptan** | antimigraine | Clinician-recommended relevance |
| zolmitriptan | 135775 | **triptan** | antimigraine | Clinician-recommended relevance |
| almotriptan | 279645 | **triptan** | antimigraine | Clinician-recommended relevance |
| rizatriptan | 88014 | **triptan** | antimigraine | Clinician-recommended relevance |
| eletriptan | 231049 | **triptan** | antimigraine | Clinician-recommended relevance |
| frovatriptan | 228783 | **triptan** | antimigraine | Clinician-recommended relevance |
| ubrogepant | 2268216 | **CGRP antagonists** | antimigraine | Clinician-recommended relevance |
| rimegepant | 2282307 | **CGRP antagonists** | antimigraine | Clinician-recommended relevance |
| atogepant | 2571813 | **CGRP antagonists** | antimigraine | Clinician-recommended relevance |
| zavegepant | 2637955 | **CGRP antagonists** | antimigraine | Clinician-recommended relevance |
| erenumab | 2045613 | **CGRP antagonists** | antimigraine | Clinician-recommended relevance |
| eptinezumab | 2282660 | **CGRP antagonists** | antimigraine | Clinician-recommended relevance |
| fremanezumab | 2056691 | **CGRP antagonists** | antimigraine | Clinician-recommended relevance |
| galcanezumab | 2058846 | **CGRP antagonists** | antimigraine | Clinician-recommended relevance |
| onabotulinumtoxinA | 860189 | **Botox** | muscle relaxant | Clinician-recommended relevance |
| baclofen | 1292 | **skeletal muscle relaxant** | muscle relaxant | Clinician-recommended relevance |
| tizanidine | 57258 | **skeletal muscle relaxant** | muscle realaxant | Clinician-recommended relevance |
| celecoxib | 140587 | **COX-2 selective NSAIDS** | pain medications | Clinician-recommended relevance |
| methylprednisolone | 6902 | **corticosteroids** | anti-inflammatory | Clinician-recommended relevance |
| hydrocortisone | 5492 | **corticosteroids** | anti-inflammatory | Clinician-recommended relevance |
| prednisolone | 8638 | **corticosteroids** | anti-inflammatory | Clinician-recommended relevance |
| prednisone | 8640 | **corticosteroids** | anti-inflammatory | Clinician-recommended relevance |
| fluocinolone | 25126 | **corticosteroids** | anti-inflammatory | Clinician-recommended relevance |
| triamcinolone | 10759 | **corticosteroids** | anti-inflammatory | Clinician-recommended relevance |
| fluticasone | 41126 | **corticosteroids** | anti-inflammatory | Clinician-recommended relevance |
| beclomethasone | 1347 | **corticosteroids** | anti-inflammatory | Clinician-recommended relevance |
| betamethasone | 1514 | **corticosteroids** | anti-inflammatory | Clinician-recommended relevance |
| dexamethasone | 3264 | **corticosteroids** | anti-inflammatory | Clinician-recommended relevance |
| capsaicin | 1992 | **analgesic** | analgesic | Clinician-recommended relevance |
| lidocaine | 6387 | **anesthetic** | anesthetic | Clinician-recommended relevance |
| dronabinol | 10402 | **synthetic THC** | THC | Clinician-recommended relevance |
| nabilone | 31447 | **synthetic THC** | THC | Clinician-recommended relevance |

**Table S5**: RxNorm codes, categories, and inclusion criteria of medications.

**Prompt**

Please review the patient's medical chart as a clinician. Analyze the medical notes provided and identify the primary cause of death. If multiple causes are present, list up to three in order of relevance, separated by commas without spaces.

-	Use standardized medical diagnoses, avoiding terms that describe states or events (e.g., avoid 'asystole').

-	When appropriate, generalize specific conditions to broader diagnoses (e.g., use 'sepsis' rather than 'severe sepsis').

-	Keep diagnoses simple and consistent across all answers.

-	If the cause of death cannot be determined, return 'NA'.

-	Provide only the list of causes without any additional explanation.

Medical Notes: {note}

**Table S6**: Secure GPT-3.5 Turbo prompt for potential causes of death identification

|  | **Missing** | **Overall**  **(n=14,107)** | **new user**  **(n=9444)** | **consistent user**  **(n=4663)** | **P** |
| --- | --- | --- | --- | --- | --- |
| **Comorbid conditions** |  |  |  |  |  |
| **myocardial infarction, n (%)** | 0 | 1863 (13.2) | 1031 (10.9) | 832 (17.9) | <0.001 |
| **congestive heart failure, n (%)** | 0 | 2903 (20.6) | 1632 (17.3) | 1271 (27.3) | <0.001 |
| **peripheral vascular disease, n (%)** | 0 | 2504 (17.8) | 1332 (14.1) | 1172 (25.2) | <0.001 |
| **cerebrovascular disease, n (%)** | 0 | 3439 (24.4) | 2122 (22.5) | 1317 (28.3) | <0.001 |
| **chronic pulmonary disease, n (%)** | 0 | 2862 (20.3) | 1521 (16.1) | 1341 (28.8) | <0.001 |
| **rheumatologic disease, n (%)** | 0 | 583 (4.1) | 286 (3.0) | 297 (6.4) | <0.001 |
| **peptic ulcer, n (%)** | 0 | 424 (3.0) | 194 (2.1) | 230 (4.9) | <0.001 |
| **hemiplegia/paraplegia, n (%)** | 0 | 480 (3.4) | 281 (3.0) | 199 (4.3) | <0.001 |
| **prediabetes, n (%)** | 0 | 835 (5.9) | 494 (5.2) | 341 (7.3) | <0.001 |
| **diabetes without complications, n (%)** | 0 | 3099 (22.0) | 1754 (18.6) | 1345 (28.9) | <0.001 |
| **diabetes with chronic complications, n (%)** | 0 | 1450 (10.3) | 785 (8.3) | 665 (14.3) | <0.001 |
| **mild liver diseases, n (%)** | 0 | 1094 (7.8) | 525 (5.6) | 569 (12.2) | <0.001 |
| **moderate severe liver diseases,**  **n (%)** | 0 | 220 (1.6) | 91 (1.0) | 129 (2.8) | <0.001 |
| **renal diseases, n (%)** | 0 | 3822 (27.1) | 2188 (23.2) | 1634 (35.1) | <0.001 |
| **any malignancy**  **(tumor/leukemia/lymphoma),**  **n (%)** | 0 | 2793 (19.8) | 1470 (15.6) | 1323 (28.5) | <0.001 |
| **metastatic solid tumor, n (%)** | 0 | 724 (5.1) | 308 (3.3) | 416 (8.9) | <0.001 |
| **cognitive decline, n (%)** | 0 | 1458 (10.4) | 938 (9.9) | 520 (11.2) | 0.03 |
| **hypertension, n (%)** | 0 | 8839 (62.8) | 5392 (57.2) | 3447 (74.1) | <0.001 |
| **hyperlipidemia, n (%)** | 0 | 6983 (49.6) | 4221 (44.8) | 2762 (59.4) | <0.001 |
| **excessive alcohol, n (%)** | 0 | 521 (3.7) | 268 (2.8) | 253 (5.4) | <0.001 |
| **atherosclerosis, n (%)** | 0 | 3616 (25.7) | 2040 (21.6) | 1576 (33.9) | <0.001 |
| **hypercholesterolemia, n (%)** | 0 | 1413 (10.0) | 830 (8.8) | 583 (12.5) | <0.001 |
| **atrial fibrillation, n (%)** | 0 | 3326 (23.6) | 2035 (21.6) | 1291 (27.8) | <0.001 |
| **hearing loss, n (%)** | 0 | 2112 (15.0) | 1300 (13.8) | 812 (17.5) | <0.001 |
| **sleep apnea, n (%)** | 0 | 2164 (15.4) | 1209 (12.8) | 955 (20.5) | <0.001 |
| **delirium, n (%)** | 0 | 1633 (11.6) | 905 (9.6) | 728 (15.7) | <0.001 |
| **depression, n (%)** | 0 | 2975 (21.1) | 1642 (17.4) | 1333 (28.7) | <0.001 |
| **schizophrenia/non-mood psychotic disorder, n (%)** | 0 | 649 (4.6) | 384 (4.1) | 265 (5.7) | <0.001 |
| **mood disorder, n (%)** | 0 | 3310 (23.5) | 1844 (19.6) | 1466 (31.5) | <0.001 |
| **anxiety/non-psychotic mental disorder, n (%)** | 0 | 3153 (22.4) | 1725 (18.3) | 1428 (30.7) | <0.001 |
| **behavioral syndromes associated with physiological disturbances and physical factors, n(%)** | 0 | 515 (3.7) | 254 (2.7) | 261 (5.6) | <0.001 |
| **disorders of adult personality and behavior, n (%)** | 0 | 126 (0.9) | 79 (0.8) | 47 (1.0) | 0.35 |
| **developmental disorder, n (%)** | 0 | 105 (0.7) | 60 (0.6) | 45 (1.0) | 0.04 |
| **behavioral and emotional disorder with onset in early ages, n (%)** | 0 | 126 (0.9) | 69 (0.7) | 57 (1.2) | 0.01 |
| **Medications** |  |  |  |  |  |
| **COX-2 selective NSAIDs, n (%** | 0 | 530 (3.8) | 219 (2.3) | 311 (6.7) | <0.001 |
| **non-COX-2 selective NSAIDs, n (%)** | 0 | 7139 (50.7) | 4238 (44.9) | 2901 (62.4) | <0.001 |
| **GI antispasmodics, n (%)** | 0 | 1653 (11.7) | 830 (8.8) | 823 (17.7) | <0.001 |
| **SGLT2 inhibitors, n (%)** | 0 | 247 (1.8) | 133 (1.4) | 114 (2.5) | <0.001 |
| **SNRIs, n (%)** | 0 | 1158 (8.2) | 568 (6.0) | 590 (12.7) | <0.001 |
| **SSRIs, n (%)** | 0 | 4445 (31.6) | 2698 (28.6) | 1747 (37.6) | <0.001 |
| **TCAs, n (%)** | 0 | 477 (3.4) | 234 (2.5) | 243 (5.2) | <0.001 |
| **TeCA, n (%)** | 0 | 973 (6.9) | 579 (6.1) | 394 (8.5) | <0.001 |
| **acetaminophen, n (%)** | 0 | 10485 (74.5) | 6321 (67.0) | 4164 (89.6) | <0.001 |
| **acetylcholinesterase inhibitors, n (%)** | 0 | 3454 (24.5) | 2672 (28.3) | 782 (16.8) | <0.001 |
| **amiodarone, n (%)** | 0 | 621 (4.4) | 311 (3.3) | 310 (6.7) | <0.001 |
| **androgens, n (%)** | 0 | 122 (0.9) | 72 (0.8) | 50 (1.1) | 0.075 |
| **anticonvulsants, n (%)** | 0 | 4094 (29.1) | 2149 (22.8) | 1945 (41.8) | <0.001 |
| **antipsychotics, n (%)** | 0 | 2805 (19.9) | 1735 (18.4) | 1070 (23.0) | <0.001 |
| **aspirin, n (%)** | 0 | 5311 (37.7) | 3286 (34.8) | 2025 (43.6) | <0.001 |
| **barbiturates, n (%)** | 0 | 231 (1.6) | 117 (1.2) | 114 (2.5) | <0.001 |
| **benzodiazepines, n (%)** | 0 | 5670 (40.3) | 2943 (31.2) | 2727 (58.7) | <0.001 |
| **blood thinners, n (%)** | 0 | 115 (0.8) | 59 (0.6) | 56 (1.2) | <0.001 |
| **botox, n (%)** | 0 | 136 (1.0) | 63 (0.7) | 73 (1.6) | <0.001 |
| **calcium channel blockers, n (%)** | 0 | 315 (2.2) | 148 (1.6) | 167 (3.6) | <0.001 |
| **central alpha agonists, n (%)** | 0 | 498 (3.5) | 240 (2.5) | 258 (5.5) | <0.001 |
| **corticosteroids, n (%)** | 0 | 5994 (42.6) | 3197 (33.9) | 2797 (60.2) | <0.001 |
| **desmopressin, n (%)** | 0 | 98 (0.7) | 43 (0.5) | 55 (1.2) | <0.001 |
| **digoxine, n (%)** | 0 | 477 (3.4) | 282 (3.0) | 195 (4.2) | <0.001 |
| **diuretics, n (%)** | 0 | 4813 (34.2) | 2785 (29.5) | 2028 (43.6) | <0.001 |
| **estrogens, n (%)** | 0 | 498 (3.5) | 298 (3.2) | 200 (4.3) | 0.001 |
| **first-generation antihistamines, n (%)** | 0 | 3890 (27.6) | 1782 (18.9) | 2108 (45.3) | <0.001 |
| **insulin, n (%)** | 0 | 2908 (20.7) | 1648 (17.5) | 1260 (27.1) | <0.001 |
| **megestrol, n (%)** | 0 | 173 (1.2) | 89 (0.9) | 84 (1.8) | <0.001 |
| **metoclopramide, n (%)** | 0 | 2817 (20.0) | 1415 (15.0) | 1402 (30.2) | <0.001 |
| **mineral oil, n (%)** | 0 | 1602 (11.4) | 869 (9.2) | 733 (15.8) | <0.001 |
| **nifedipine, n (%)** | 0 | 166 (1.2) | 95 (1.0) | 71 (1.5) | 0.01 |
| **nitrofuratoin, n (%)** | 0 | 812 (5.8) | 430 (4.6) | 382 (8.2) | <0.001 |
| **non-selective peripheral alpha-1-blockers, n (%)** | 0 | 364 (2.6) | 229 (2.4) | 135 (2.9) | 0.11 |
| **nonbenzodiazepine benzodiazepine receptor agonist hypnotics, n (%)** | 0 | 1386 (9.8) | 672 (7.1) | 714 (15.4) | <0.001 |
| **phenyltriazine, n (%)** | 0 | 254 (1.8) | 160 (1.7) | 94 (2.0) | 0.20 |
| **proton pump inhibitors, n (%)** | 0 | 4759 (33.8) | 2615 (27.7) | 2144 (46.1) | <0.001 |
| **rivaroxaban, n (%)** | 0 | 529 (3.8) | 303 (3.2) | 226 (4.9) | <0.001 |
| **skeletal muscle relaxant, n (%)** | 0 | 954 (6.8) | 398 (4.2) | 556 (12.0) | <0.001 |
| **sulfonylureas, n (%)** | 0 | 750 (5.3) | 426 (4.5) | 324 (7.0) | <0.001 |
| **anesthetic, n (%)** | 0 | 6991 (49.7) | 3875 (41.1) | 3116 (67.0) | <0.001 |
| **triptan, n (%)** | 0 | 128 (0.9) | 61 (0.6) | 67 (1.4) | <0.001 |
| **warfarin, n (%)** | 0 | 802 (5.7) | 464 (4.9) | 338 (7.3) | <0.001 |

**Table S7:** Descriptive statistics of opioid-exposed patients, comparing new users and consistent users on comorbidities and medication exposure

Categorical variables were compared using the chi-squared test. Continuous variables were evaluated using Welch’s t-test. Benjamini-Hochberg method was used to correct for multiple testing errors. All analyses were conducted using the PyPI package tableone (v.0.9.1)^17^. Variables that contained cells with fewer than 20 cases were dropped to protect patient privacy. Variables are defined in Table S3.

|  | **Missing** | **Overall (n=27,759)** | **exposed (n=14,107)** | **unexposed (n=13,652)** | **P** |
| --- | --- | --- | --- | --- | --- |
| **Exposure group, n (%)** |  |  |  |  |  |
| **consistent user** | 0 | 4663 (16.8) | 4663 (33.1) | 0 (0) | <0.001 |
| **new user** |  | 9444 (34.0) | 9444 (66.9) | 0 (0) |  |
| **control** |  | 11973 (43.1) | 0 (0) | 11973 (87.7) |  |
| **discontinued** |  | 1679 (6.0) | 0 (0) | 1679 (12.3) |  |
| **Dementia category** |  |  |  |  |  |
| **Mild cognitive impairment, n (%)** |  | 10229 (36.8) | 5213 (37.0) | 5016 (36.7) | 0.72 |
| **Alzheimer’s disease, n (%)** |  | 6639 (23.9) | 3160 (22.4) | 3479 (25.5) | <0.001 |
| **Vascular dementia, n (%)** |  | 2351 (8.5) | 1319 (9.3) | 1032 (7.6) | <0.001 |
| **Other/unspecified dementia, n (%)** |  | 20301 (73.1) | 10637 (75.4) | 9664 (70.8) | <0.001 |
| **Sex, n (%)** |  |  |  |  |  |
| **Female** | 2 | 15838 (57.1) | 7904 (56.0) | 7934 (58.1) | <0.001 |
| **Male** |  | 11919 (42.9) | 6202 (44.0) | 5717 (41.9) |  |
| **Age at diagnosis, median [Q1,Q3]** | 0 | 81.0 [73.0,87.0] | 81.0 [73.0,87.0] | 81.0 [74.0,87.0] | <0.001 |
| **Age group, n (%)** |  |  |  |  |  |
| **50-59** | 0 | 1258 (4.5) | 695 (4.9) | 563 (4.1) | <0.001 |
| **60-69** |  | 3141 (11.3) | 1720 (12.2) | 1421 (10.4) |  |
| **70-79** |  | 8139 (29.3) | 4080 (28.9) | 4059 (29.7) |  |
| **80-89** |  | 10777 (38.8) | 5499 (39.0) | 5278 (38.7) |  |
| **90-100** |  | 4444 (16.0) | 2113 (15.0) | 2331 (17.1) |  |
| **Race, n (%)** |  |  |  |  |  |
| **White** | 137 | 17464 (63.2) | 9021 (64.2) | 8443 (62.2) | <0.001 |
| **Asian** |  | 4065 (14.7) | 2019 (14.4) | 2046 (15.1) |  |
| **Black** |  | 1378 (5.0) | 725 (5.2) | 653 (4.8) |  |
| **Native American/Hawaiian/Pacific Islander** |  | 183 (0.7) | 117 (0.8) | 66 (0.5) |  |
| **Unknown/Refused** |  | 916 (3.3) | 265 (1.9) | 651 (4.8) |  |
| **Other** |  | 3616 (13.1) | 1903 (13.5) | 1713 (12.6) |  |
| **Ethnicity, n (%)** |  |  |  |  |  |
| **Hispanic/Latino** | 67 | 2370 (8.6) | 1326 (9.4) | 1044 (7.7) | <0.001 |
| **Non-Hispanic/Non-Latino** |  | 24053 (86.9) | 12402 (88.0) | 11651 (85.7) |  |
| **Unknown/Refused** |  | 1269 (4.6) | 366 (2.6) | 903 (6.6) |  |
| **Death, n (%)** |  | 10998 (39.6) | 6104 (43.3) | 4894 (35.8) | <0.001 |
| **Death from diagnosis, median [Q1,Q3]** | 16761 | 849.0 [269.0,1682.8] | 829.5 [233.0,1713.2] | 872.5 [312.0,1641.8] | 0.053 |

**Table S8:** Descriptive statistics of the dementia/mild cognitive impairment cohort based on post-diagnosis opioid exposure status

Categorical variables were compared using the chi-squared test. Continuous variables were evaluated using Welch’s t-test. Benjamini-Hochberg method was used to correct for multiple testing errors. All analyses were conducted using the PyPI package tableone (v.0.9.1)^17^. Variables that contained cells with fewer than 20 cases were dropped to protect patient privacy. Variables are defined in Table S3.

|  | | **Missing** | **Overall (n=113,343)** | **new user (n=77,168)** | **consistent user**  **(n=36175)** | ***P*** |
| --- | --- | --- | --- | --- | --- | --- |
| **Dementia category** | | **0** |  |  |  |  |
| **Mild cognitive impairment, n (%)** | |  | 30312 (26.7) | 19941 (25.8) | 10371 (28.7) | <0.001 |
| **Alzheimer’s disease, n (%)** | |  | 39758 (35.1) | 29369 (38.1) | 10389 (28.7) | <0.001 |
| **Vascular dementia, n (%)** | |  | 19836 (17.5) | 13279 (17.2) | 6557 (18.1) | <0.001 |
| **Frontotemporal dementia, n (%)** | |  | 2077 (1.8) | 1563 (2.0) | 514 (1.4) |  |
| **Lewy body dementia, n(%)** | |  | 2570 (2.3) | 1852 (2.4) | 718 (2.0) |  |
| **Other/Unspecified dementia, n (%)** | |  | 77823 (68.7) | 53757 (69.7) | 24066 (66.5) | <0.001 |
| **Patient characteristics** | |  |  |  |  |  |
| **Sex** | | 2 |  |  |  | <0.001 |
| Female | |  | 68809 (60.7) | 46489 (60.2) | 22320 (61.7) |  |
| Male | |  | 44532 (39.3) | 30677 (39.8) | 13855 (38.3) |  |
| **Age at diagnosis, median [Q1,Q3]** | | 0 | 81.0 [74.0,87.0] | 80.0 [73.0,87.0] | 81.0 [74.0,87.0] | <0.001 |
| **Age group, n (%)** | | 0 |  |  |  | <0.001 |
| 50-59 | |  | 4567 (4.0) | 2893 (3.7) | 1674 (4.6) |  |
| 60-69 | |  | 13255 (11.7) | 8561 (11.1) | 4694 (13.0) |  |
| 70-79 | |  | 32210 (28.4) | 21613 (28.0) | 10579 (29.3) |  |
| 80-89 | |  | 44102 (38.9) | 30649 (39.7) | 13453 (37.2) |  |
| 90-100 | |  | 19209 (16.9) | 13452 (17.4) | 5757 (15.9) |  |
| **Race, n (%)** | | 0 |  |  |  | <0.001 |
| White | |  | 93131 (82.2) | 62781 (81.4) | 30350 (83.9) |  |
| Black | |  | 3468 (3.1) | 2256 (2.9) | 1212 (3.4) |  |
| Asian | |  | 5022 (4.4) | 3841 (5.0) | 1181 (3.3) |  |
| Native American/Hawaiian/Pacific Islander | |  | 1140 (1.0) | 731 (0.9) | 409 (1.1) |  |
| Unknown/Refused/Declined | |  | 4529 (4.0) | 3408 (4.4) | 1121 (3.1) |  |
| Other | |  | 6053 (5.3) | 4151 (5.4) | 1902 (5.3) |  |
| **Ethnicity, n (%)** |  | 0 |  |  |  | <0.001 |
| Hispanic/Latino |  |  | 6296 (5.6) | 4283 (5.6) | 2013 (5.6) |  |
| Non-Hispanic/Latino |  |  | 103405 (91.2) | 70097 (90.8) | 33308 (92.1) |  |
| Unknown/Refused/Declined |  |  | 3642 (2.6) | 2788 (3.6) | 854 (2.4) |  |
| **Insurance, n (%)** |  | 3095 |  |  |  | 0.78 |
| Medicare/Medicaid |  |  | 105016 (92.7) | 70444 (91.3) | 2470 (53.6) |  |
| Private |  |  | 5219 (4.6) | 4009 (5.2) | 1468 (31.9) |  |
| Other |  |  | N/A | N/A | N/A |  |
| **BMI, median [Q1,Q3]** | | 9278 | 25.4 [22.2,29.4] | 25.9 [22.5,30.3] | 25.2 [22.0,29.1] | <0.001 |
| **BMI category, n (%)** | | 9278 |  |  |  | <0.001 |
| Under weight | |  | 5839 (5.2) | 4009 (5.2) | 1830 (5.1) |  |
| Normal weight | |  | 41763 (36.8) | 28930 (37.5) | 12833 (35.5) |  |
| Over weight | |  | 32527 (28.7) | 21801 (28.3) | 10726 (29.7) |  |
| Obese | |  | 15183 (13.4) | 9622 (12.5) | 5561 (15.4) |  |
| Severely obese | |  | 5771 (5.1) | 3389 (4.4) | 2382 (6.6) |  |
| **Death outcome** |  |  |  |  |  |  |
| **Days to death from first opioid exposure, mean (SD)** | | 70200 | 634.0 (764.2) | 617.7 (772.0) | 669.1 (745.9) | <0.001 |
| **Death within 14 days after first opioid exposure, n (%)** | | 0 | 7022 (6.2) | 5401 (7.0) | 1621 (4.5) | <0.001 |
| **Death within 60 days after first opioid exposure, n (%)** | | 0 | 12593 (11.1) | 9300 (12.1) | 3293 (9.1) | <0.001 |
| **Death within 180 days after first opioid exposure, n (%)** | | 0 | 18024 (15.9) | 12970 (16.8) | 5054 (14.0) | <0.001 |

**Table S9 Descriptive statistics of the validation analytic cohort**

Categorical variables were compared using the chi-squared test. Continuous variables were evaluated using Welch’s t-test. The Benjamini-Hochberg method was used to correct for multiple testing errors. All analyses were conducted using the PyPI package tableone (v.0.9.1)^17^. Variables that contained cells with fewer than 20 cases were dropped to protect patient privacy. Variables are defined in Table S3.

| **Note** | **Extracted principal diagnosis at time of death** | **Cause 1** | **Cause 2** | **Cause 3** | **Extracted principal diagnosis health condition category** | **Causes health condition category** |
| --- | --- | --- | --- | --- | --- | --- |
| **Principle Diagnosis at Time of Death:** AICD lead infection  **Relevant Clinical Course:**  **-** Male with ischemic cardiomyopathy and previous AICD placement was admitted due to septic shock and septic pulmonary emboli from an infected AICD lead.  - The patient presented with hypotension, hypoxia, and a large vegetation on the right ventricular pacemaker lead.  - The patient experienced worsening respiratory failure, bradycardia, and a ventricular tachycardia arrest, leading to multiple resuscitations.  **Date of Death:** [deidentified]  **Time of Death:** [deidentified]  **Comorbidities and Complications:**  -Septic shock due to coag-negative staph and Candida albican Elevated liver enzymes  - Cardiac/pericardial tamponade  - Bacteremia  - Pulmonary embolus  - Decubitus ulcer (stage 3)  - Anemia  - Septic embolism | AICD lead infection | Septic shock | Cardiac tamponade | Pulmonary embolus | Infection, cardiovascular | Sepsis, cardiovascular, respiratory |

**Table S9** Notes, GPT Responses, and Health Condition Categories of Mismatches Between Specified Principal Diagnoses at the Time of Death and GPT Responses.

Clinical free-text notes were preprocessed using Stanford’s secure GPT-4o model to deidentify protected health information and convert the text into a format suitable for protecting patient and clinician privacy while increasing readability. The prompt was “Please analyze the following clinical text notes and: 1. Remove all identifiable information, including names, exact dates, locations, or any other PHI. 2. Replace any mentions of the exact time of death with deidentified placeholders (e.g., "Time of death: [deidentified]"). 3. Retain the patient's age but ensure no other potentially identifying details are included. 4. Extract only the content related to death, such as causes, circumstances, or timeframes. 5. Summarize the extracted information in a concise, non-identifiable format suitable for inclusion in a publicly available manuscript. Ensure the final summary adheres to data privacy regulations like HIPAA and is completely deidentified. Here’s the text: [clinical text]”

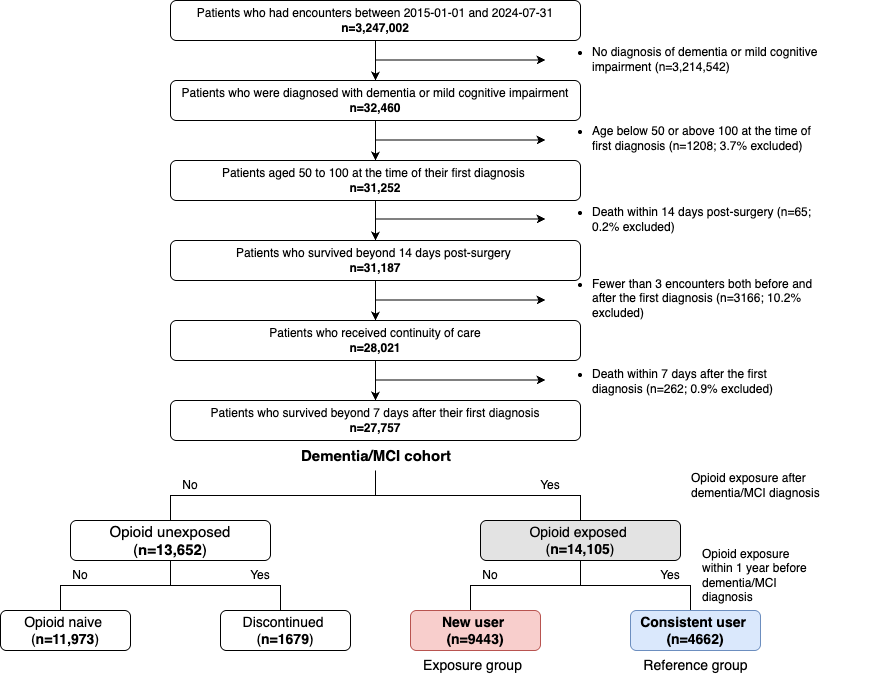

**Figure S1 Flowchart of cohort and exposure group selection in the validation cohort**

Descriptive statistics of the dementia/MCI cohort comparing opioid-exposed and unexposed groups are presented in Table S8. Descriptive statistics of the opioid-exposed cohort comparing new users and consistent users are presented in Table 1 and Table S7.

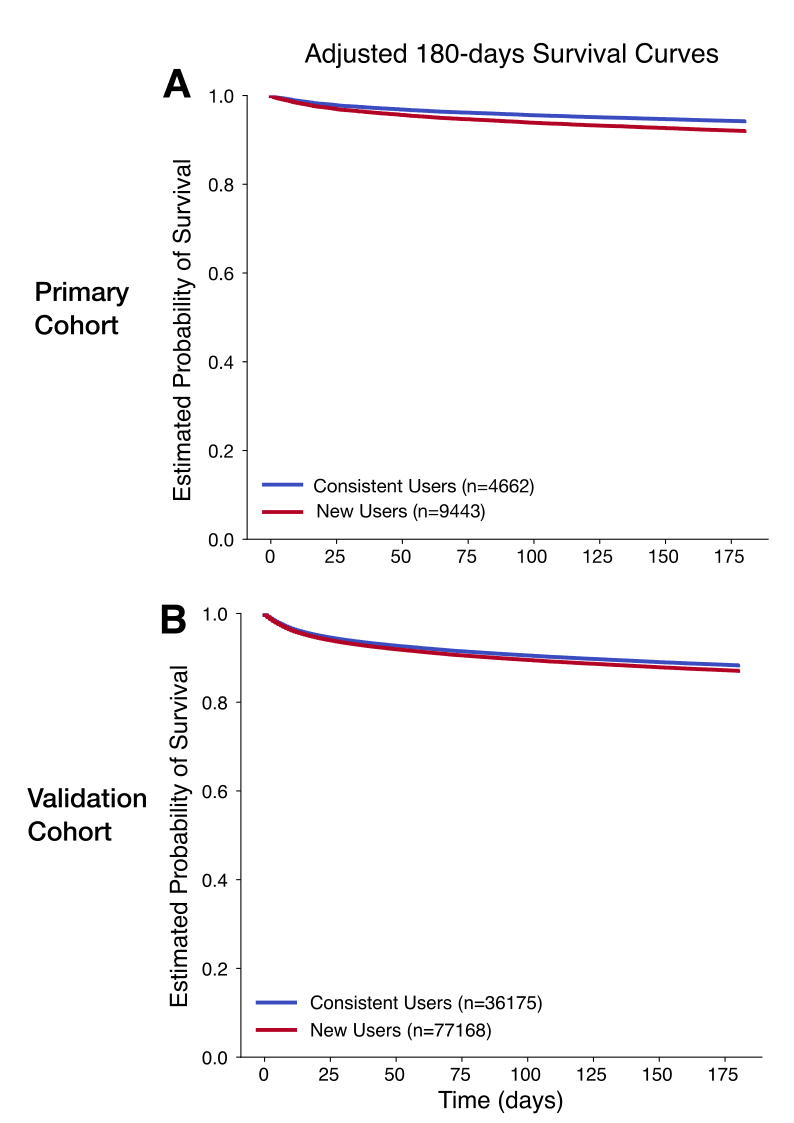

**Figure S2: 180-day survival probability curves comparing new users and consistent users in primary and validation cohorts**

A. 180-day survival probability curve comparing new users and consistent users in primary cohort. B. 180-day survival probability curve comparing new users and consistent users in validation cohort. In primary cohort, new opioid users (n=9443) exhibited a higher adjusted hazard ratio (aHR) for 180-day survival following opioid initiation, with an aHR of 1.40 (95% CI: [1.25, 1.56], P < 0.0001). The predictive model demonstrated strong performance, achieving a concordance index of 0.73. In validation cohort, new opioid users (n=77,168) exhibited a higher aHR for 180-day survival following opioid initiation, with an aHR of 1.11 (95% CI: [1.07, 1.15], P < 0.0001). The predictive model demonstrated strong performance, achieving a concordance index of 0.77. Hazard ratios were estimated using a Cox proportional hazards model, adjusting for age at diagnosis, race, ethnicity, body mass index (BMI), insurance status, comorbidities, and medication exposures. Comprehensive lists of comorbidities and medications included in the model are provided in Tables S4 and S5. Variables with cases <5% of analytic cohort were excluded from the analysis. Statistical significance levels are denoted as follows: +: P < 0.1, *: P < 0.05, **: P < 0.01, ***: P < 0.001, ****: P < 0.0001.

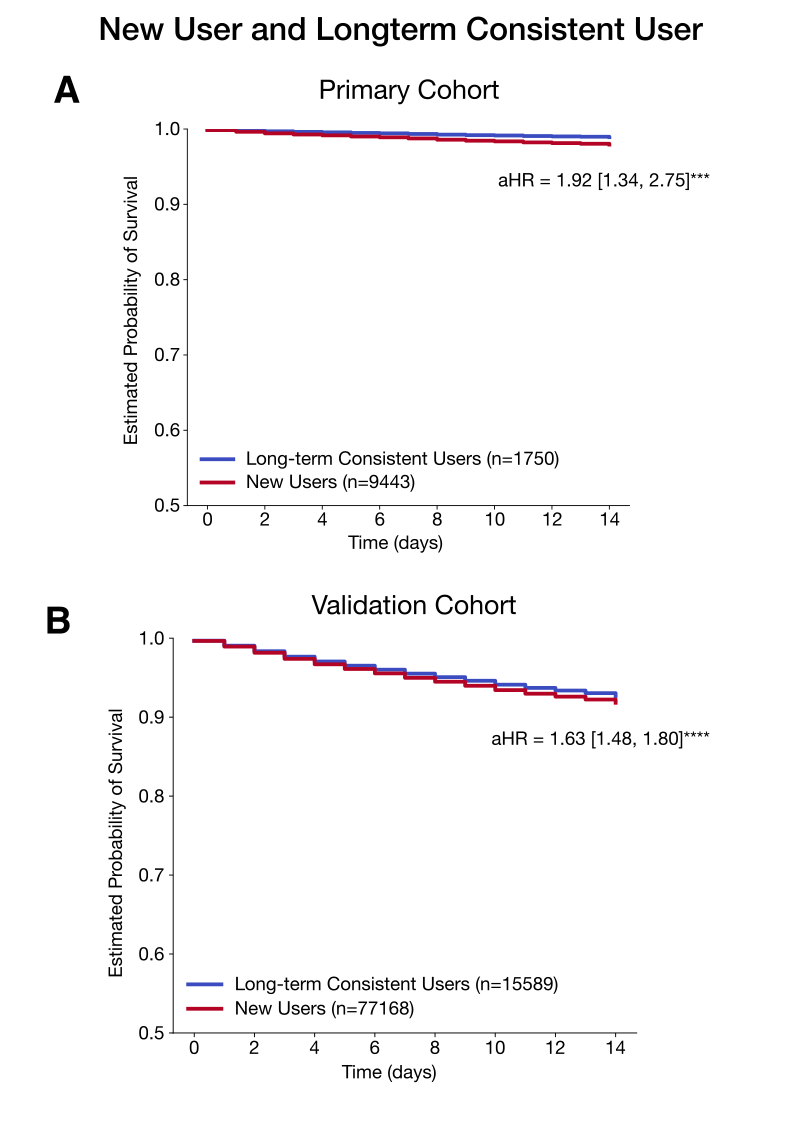

**Figure S3: Sensitivity analysis comparing first-time opioid users to long-term consistent users in primary and validation cohort**

A. sensitivity analysis comparing first-time opioid users to individuals who had used opioids consistently for at least 90 days during the pre-diagnosis observation period (long-term consistent users) in the primary cohort. B. sensitivity analysis comparing first-time opioid users to long-term consistent users in the validation cohort. The association between opioid initiation and short-term mortality remained robust, with an adjusted hazard ratio (aHR) of 1.92 [1.28–2.65] (P<0.001) in primary cohort and aHR of 1.63 [1.48, 1.80] (P<0.0001) in validation cohort. Significance levels: +: P < 0.1, *: P < 0.05, **: P < 0.01, ***: P < 0.001, ****: P < 0.0001.

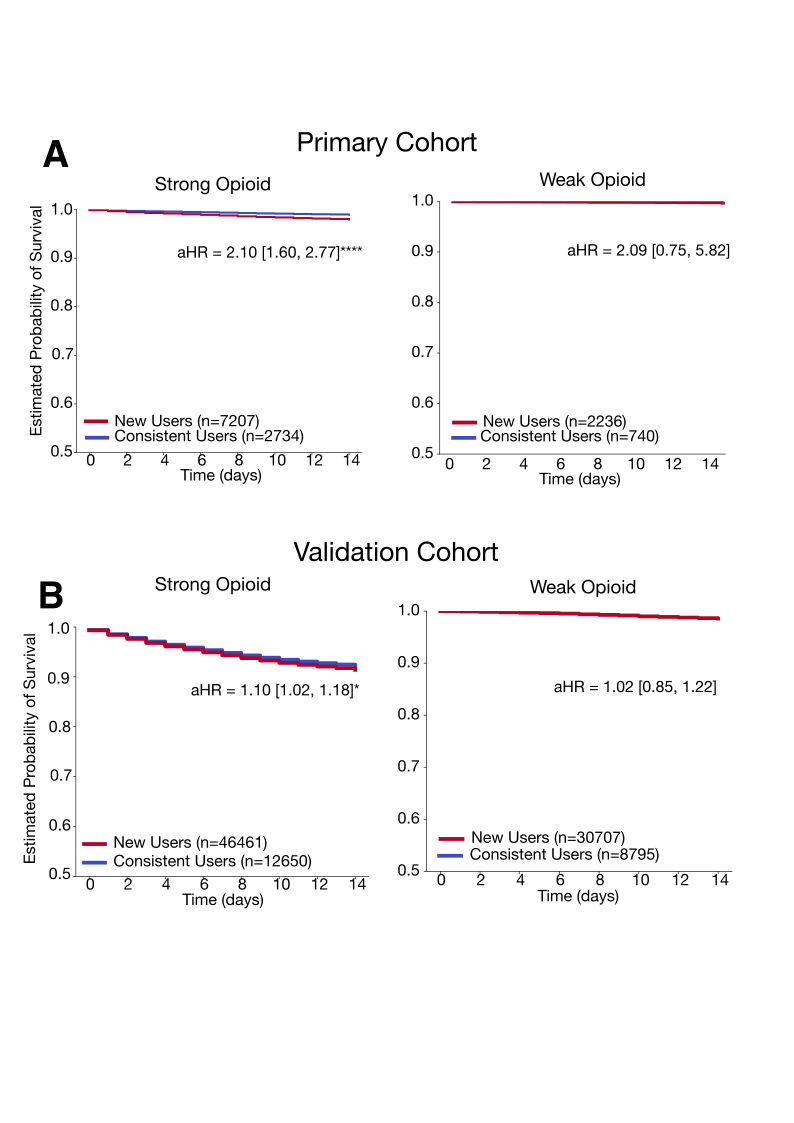

**Figure S4: Strong opioid-specific and weak-opioid specific 14-day survival probability curve comparing new users and consistent users in primary and validation cohort**

A. Strong opioid-specific and weak opioid-specific 14 day survival probability curve comparing new users and consistent users in primary cohort B. Strong opioid-specific and weak opioid-specific 14 day survival probability curve comparint new users and consistent users in validation cohort. These plots show the results of subgroup analysis categorizing opioid exposures into strong and weak opioids. Strong opioids included buprenorphine, fentanyl, hydromorphone, oxycodone, morphine, methadone, and meperidine. Weak opioids included hydrocodone, codeine, and tramadol. Initiation of strong opioids was associated with an increased risk of short-term mortality, with an adjusted hazard ratio (aHR) of 2.10 [1.60, 2.77] (P<0.0001) in primary cohort and 1.10 [1.02, 1.08] (P<0.05) in validation cohort. In contrast, initiation of weak opioids showed no association with short-term mortality risk in both cohorts. Significance levels: +: P < 0.1, *: P < 0.05, **: P < 0.01, ***: P < 0.001, ****: P < 0.0001.

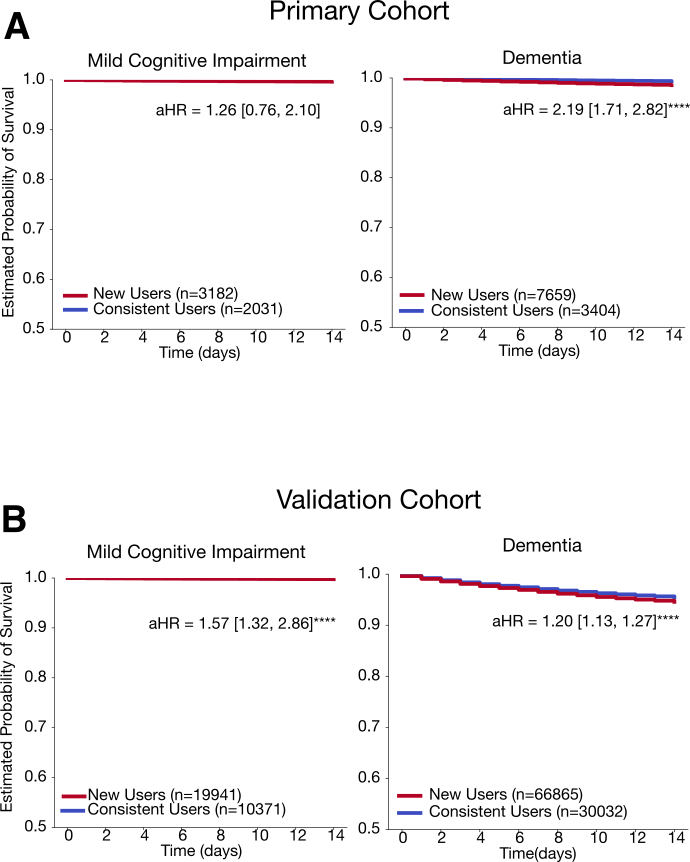

**Figure S5: MCI-specific and dementia-specific 14-day survival probability curve comparing new users and consistents users in primary and validation cohort**

A. MCI-specific and dementia-specific 14 day survival probability curve comparing new users and consistent users in primary cohort B. MCI-specific and dementia-specific 14 day survival probability curve comparint new users and consistent users in validation cohort. These plots show the results of subgroup analysis categorizing patients into MCI and dementia subgroup. Patients who had any diagnosis of MCI was included in the MCI subgroup. Patients who had any diagnosis of dementia was included in the dementia subgroup. These MCI and subgroups are not mutually exclusive. Initiation of opioids was associated with an increased risk of short-term mortality, with an adjusted hazard ratio (aHR) of 2.19 [1.71, 2.82] (P<0.0001) in dementia subgroup of primary cohort, with aHR of 1.57 [1.32, 2.86] (P<0.0001) in MCI subgroup of validation cohort, and with aHR of 1.20 [1.13,2.27] (P<0.0001) in validation cohort. The validation cohort showed larger HR in the MCI group though dementia subgroup had higher absolute mortality. Significance levels: +: P < 0.1, *: P < 0.05, **: P < 0.01, ***: P < 0.001, ****: P < 0.0001.

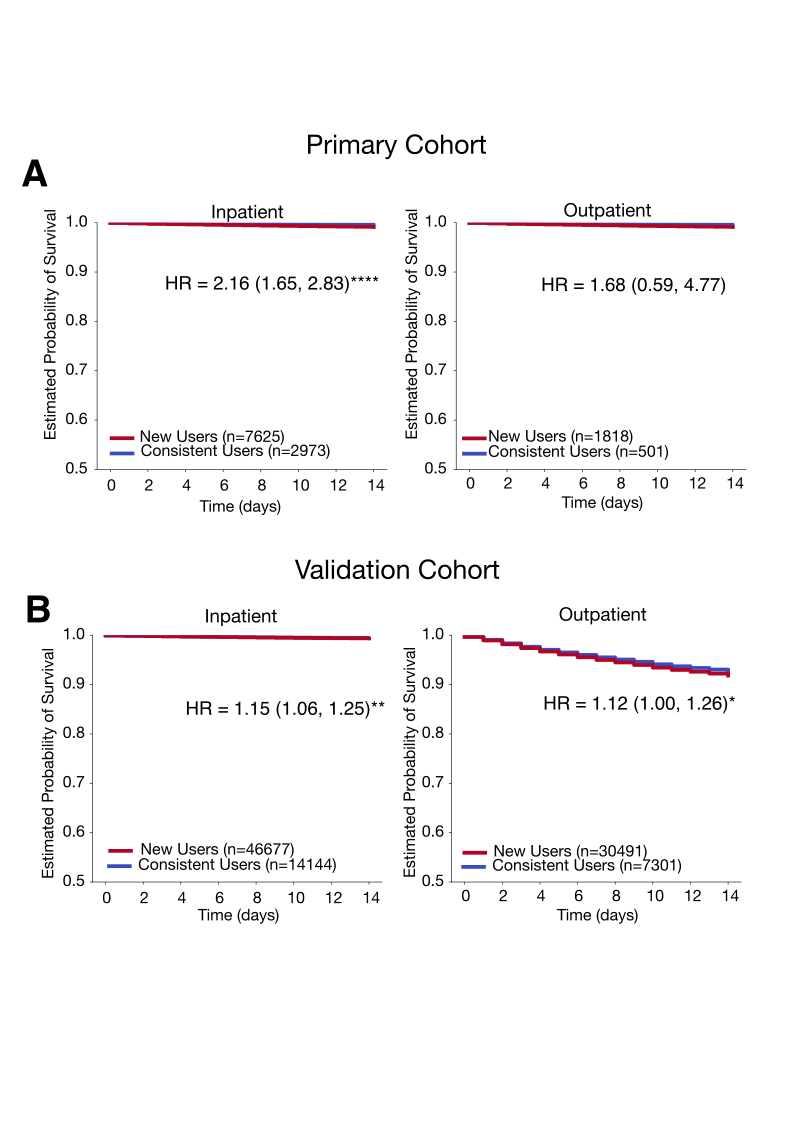

**Figure S6: Inpatient-specific and outpatient-specific 14-day survival probability curve comparing new users and consistents users in primary and validation cohort**

A. Inpatient-specific and outpatient-specific 14 day survival probability curve comparing new users and consistent users in primary cohort B. Inpatient-specific and outpatient-specific 14 day survival probability curve comparing new users and consistent users in validtation cohort. These plots show the results of subgroup analysis categorizing opioid medication order into inpatient and outpatient subgroup. Initiation of opioids was associated with an increased risk of short-term mortality, with an adjusted hazard ratio (aHR) of 2.16 [1.65, 2.83] (P<0.0001) in inpatient subgroup of primary cohort, with aHR of 1.15 [1.06, 1.25] (P<0.01) in inpatient subgroup of validation cohort, and with aHR of 1.12 [1.00, 1.26] (P<0.05) in validation cohort. The validation cohort showed larger HR in the inpatient group though outpatient subgroup had higher absolute mortality. Significance levels: +: P < 0.1, *: P < 0.05, **: P < 0.01, ***: P < 0.001, ****: P < 0.0001.

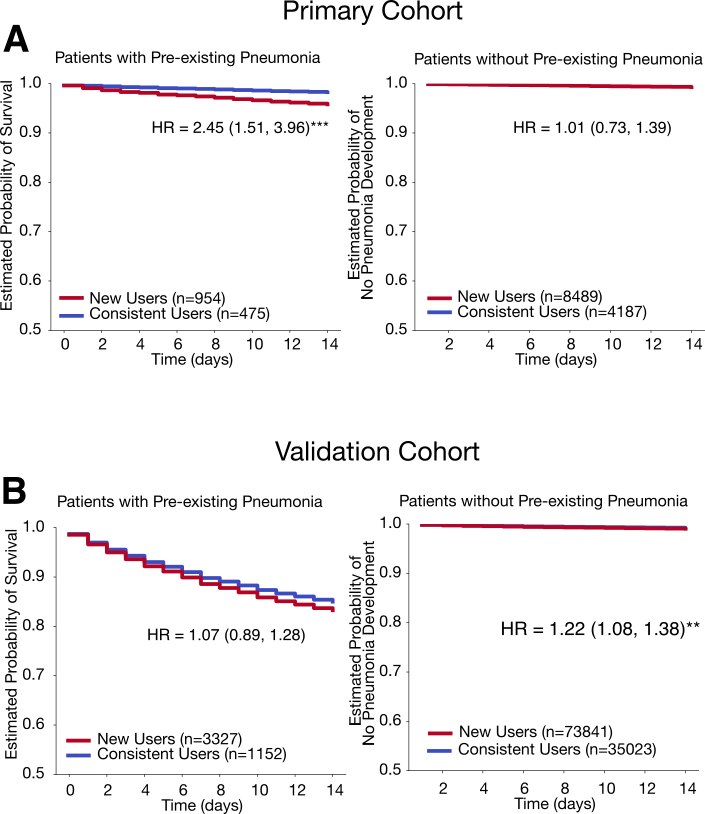

**Figure S7:Post-hoc supplementary analysis of short-term mortality associated with opioid initiation in patients with preexising pneumonia and pneumonia development risk following opioid initiation in patients without preexisting pneumonia in primary and validation cohort**

Plot A presents a sensitivity analysis of short-term mortality in patients with underlying pneumonia, showing an adjusted hazard ratio (aHR) of 2.38 (95% CI: [1.45, 3.91]). Plot B displays a sensitivity analysis examining the risk of developing pneumonia among individuals without preexisting pneumonia, with no significant association observed between opioid initiation and pneumonia risk (aHR = 1.00, 95% CI: [0.73, 1.38]). Significance levels: +: P < 0.1, *: P < 0.05, **: P < 0.01, ***: P < 0.001, ****: P < 0.0001.
